# Impact of sample clarification by size exclusion on virus detection and diversity in wastewater-based epidemiology

**DOI:** 10.1101/2022.09.25.22280344

**Authors:** Temitope O.C. Faleye, Peter Skidmore, Amir Elyaderani, Sangeet Adhikari, Nicole Kaiser, Abriana Smith, Allan Yanez, Tyler Perleberg, Erin M. Driver, Rolf U. Halden, Arvind Varsani, Matthew Scotch

## Abstract

The use of wastewater-based epidemiology (WBE) for early detection of virus circulation and response during the SARS-CoV-2 pandemic increased interest in and use of virus concentration protocols that are quick, scalable, and efficient. One such protocol involves sample clarification by size fractionation using either low-speed centrifugation to produce a clarified supernatant or membrane filtration to produce an initial filtrate depleted of solids, eukaryotes and bacterial present in wastewater (WW), followed by concentration of virus particles by ultrafiltration of the above. While this approach has been successful in identifying viruses from WW, it assumes that majority of the viruses of interest should be present in the fraction obtained by ultrafiltration of the initial filtrate, with negligible loss of viral particles and viral diversity.

We used WW samples collected in a population of ∼700,000 in southwest USA between October 2019 and March 2021, targeting three non-enveloped viruses (enteroviruses [EV], canine picornaviruses [CanPV], and human adenovirus 41 [Ad41]), to evaluate whether size fractionation of WW prior to ultrafiltration leads to appreciable differences in the virus presence and diversity determined.

We showed that virus presence or absence in WW samples in both portions (filter trapped solids [FTS] and filtrate) are not consistent with each other. We also found that in cases where virus was detected in both fractions, virus diversity (or types) captured either in FTS or filtrate were not consistent with each other. Hence, preferring one fraction of WW over the other can undermine the capacity of WBE to function as an early warning system and negatively impact the accurate representation of virus presence and diversity in a population.

## Introduction

An advantage of wastewater-based epidemiology (WBE) lies in its ability to detect presence and diversity of pathogenic human-infecting viruses in a population beyond those observed in clinical setting. Though these viruses are present at high concentrations in feces from infected individuals they are usually highly diluted in wastewater (WW) and often need to be concentrated to facilitate detectability. Over the decades, and with improving technology, different approaches have been used for concentrating viruses present in WW (please see references^1, 2, 3^ for a more detailed exploration of this topic). However, the utility of WBE for early detection of virus circulation and response during the SARS-CoV-2 pandemic increased both the interest and use of virus concentration protocols that are quick, scalable, and efficient. One such protocol involves sample clarification by size fractionation using membrane filtration to trap/exclude WW solids, eukaryotic and bacterial cells followed by concentration of suspended virus particles present in the clarified initial filtrate by ultrafiltration^4, 5, 6^. While this approach has successfully identified viruses from WW, it is predicated on the assumption that most of the viruses of interest are present in the fraction being concentrated and subsequently screened. As a corollary, the approach assumes that virus captured in filtrate only is sufficient to assess both overall virus presence and diversity. However, these two important assumptions frequently remain untested and may lead to an incomplete assessment of viral diversity in a population.

Previous studies^7,8,9,10,11^ have examined how sample clarification impacts virus abundance in both the filtrate and filter-trapped-solids usually with the goal of using one fraction to estimate what might be present or lost in the other. These studies have assumed that virus presence and diversity are one and the same and they did not examine how the clarification process impacts virus diversity in both fractions. As WBE continues to be an important tool in understanding virus presence in an early warning system capacity, it should also provide an accurate representation of viruses circulating in the population. This includes detection of all variants like immune-escape and drug-resistant mutants, and consequently provide information necessary to evaluate the utility of vaccines and chemotherapeutic agents to control these viruses or guide vaccine strain selection and antiviral drug development. Here, we evaluate whether WW clarification prior to ultrafiltration impacts virus presence and diversity in WW sample(s). Specifically, we examine virus presence and diversity in FTS and filtrate using amplicon-based high throughput sequencing of three non-enveloped viruses as case studies 1) enteroviruses (EVs), 2) canine picornaviruses (CanPV), and 3) human adenovirus 41 (Ad41).

Enteroviruses (EVs) are members of the genus *Enterovirus* in supergroup 3 (subfamily *Ensavirinae)* in the family *Picornaviridae*. EVs are viruses with non-enveloped icosahedral capsid symmetry and a diameter of 28-30nm. They have a ∼7.5kb positive-sense, single-stranded RNA genome that encodes one large polyprotein, flanked on both ends by untranslated regions (UTRs). More recently, a small ORF upstream of the large ORF has been described^12^. Over 300 EV types have been defined, grouped into 15 species, and can be typed using either complete genome, complete capsid, or complete VP1 protein gene sequence^13^. EVs infect both humans and animals, causing clinical manifestations in only about 10% of the infected. In the USA, EVs are responsible for about 15 million human infections and tens of thousands of hospitalizations annually^14^. All infected individuals shed virus in large quantities (∼10^8^ particles/gram of feces) in feces for weeks, and the virions are stable in the environment for extended periods of time^15,16,17,18,19^. Poliovirus is the most well-studied EV.

Canine picornavirus (CanPV) is a yet to be assigned member of supergroup 3 (subfamily *Ensavirinae)* in the family *Picornaviridae*. They have a ∼8kb positive-sense, single-stranded RNA genome that encodes one large polyprotein, flanked on both ends by untranslated regions (UTRs). They have been found in feces, urine, respiratory swabs, and liver (suggesting they might be capable of causing systemic infection) of dogs and red foxes in United Arab Emirates (UAE), China, Hong-Kong and Australia, and more recently in wastewater in the USA^20,21,22,23,24,25^. Very little is known about CanPV molecular diversity, and as of July 2022, less than 20 sequences were publicly available in GenBank.

Ad41 is a double-stranded DNA virus with non-enveloped capsid and etiological agent of diarrhea in children below the age of two years, often resulting in fatal systemic disseminated disease in the immunocompromised and has more recently been associated with hepatitis-of-unknown origin in the same demographic^26,27^. They are members of the genus *Mastadenovirus* in the family *Adenoviridae*. Within the seven (A-G) subgenera into which they are classified, there are over 100 types. Types 40 and 41 (Ad41) are the only members of subgenus F. Viruses in the subgenus *Adenovirus F* are transmitted via the fecal-oral route, shed in feces of infected persons and stable in wastewater for weeks^26,28^.

In this study, using these three non-enveloped viruses, we show that sample clarification by size fractionation impacts our perception of virus presence and diversity in WW. Specifically, we show that virus presence and diversity in FTS or filtrate are not usually consistent with each other. Hence, preferring one over the other can undermine the capacity of WBE to function as an early warning system and accurately represent virus presence and diversity in any population.

## Results

### Specimen characteristics

We analyzed 118 wastewater (WW) samples archived at -20°C and collected longitudinally from ten different sites in two municipalities (population; ∼700,000) in Maricopa County, Arizona, USA, between October 2019 to March 2020 (Season 1) and October 2020 to March 2021 (Season 2). For one day each month, samples from all ten sites (except March 2020 for which we had 8 sites) were recovered from the freezers and thawed overnight. Subsequently, 200 ml of wastewater from each of the ten sites was filtered using a 450 nm membrane filter. Viruses present in both filtrates and FTS were pooled independently and concentrated to 2ml using 10,000 MW cutoff centrifugal filters (please see the methods section for details). Hence, for each of the 12 months, there were two concentrates: one each for filtrate and FTS, totaling 24 concentrates.

We subjected all 24 concentrates to nucleic acid extraction using the QIAamp viral RNA MiniKit following manufacturer’s instructions. The extract was used to screen for EV, CanPV and Ad41 using a collection of nested PCR assays (Table S1 and Figure 1). Sanger sequencing confirmed a random sample of second round PCR was from respective viruses. First round amplicons of concentrates positive for the nested PCR assay were then subjected to high throughput sequencing on the Illumina platform (please see the *Methods* section for details).

**Figure 1:**
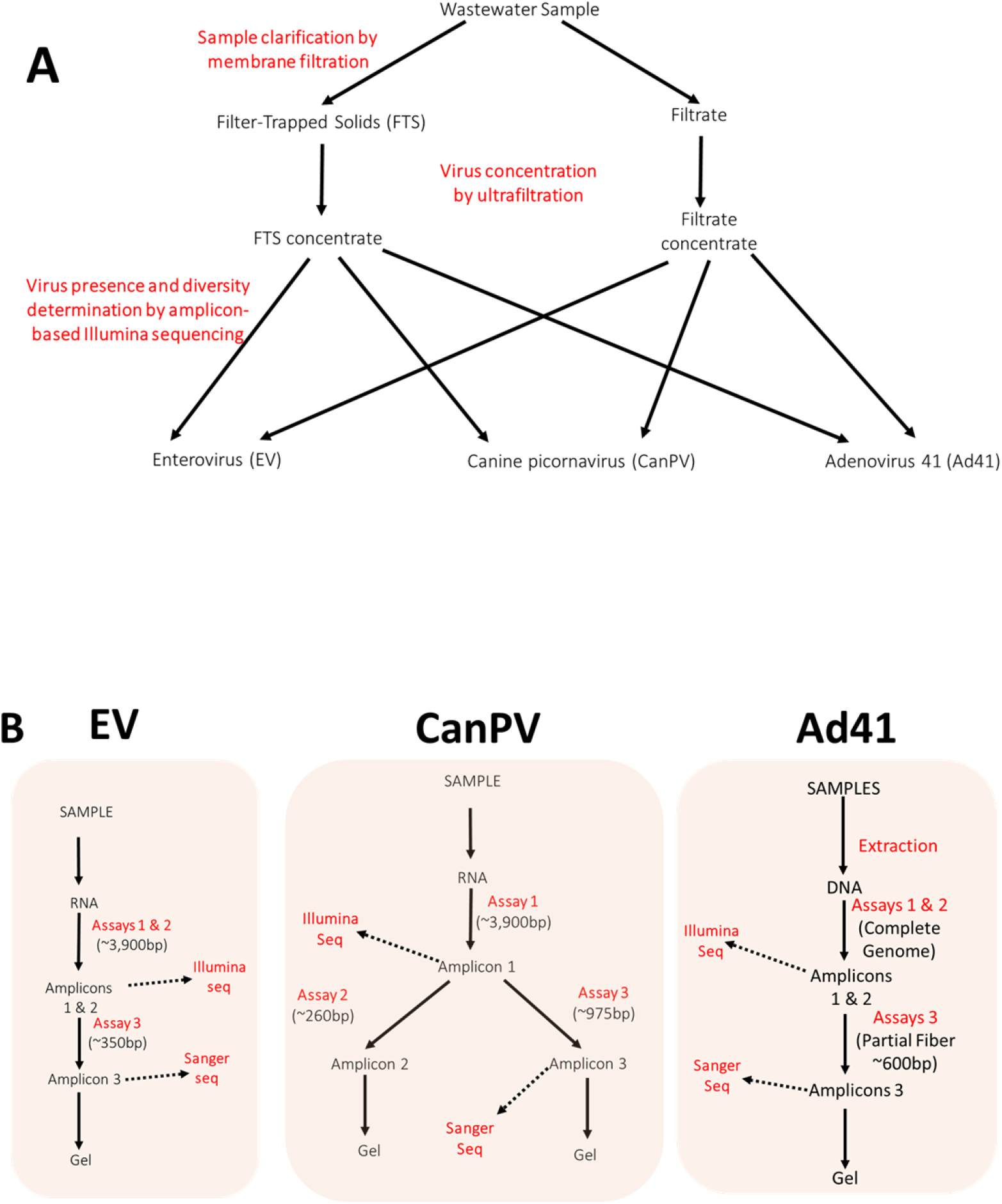
Schematic representation of A) the sample processing workflow used in this study. B) The molecular screens used in this study.

### ENTEROVIRUSES

Seventy-nine percent (19/24) of the samples were positive for the nested PCR assay suggesting the presence of EVs. Of the five negative samples, four (February 2020, October 2020, November 2020 and March 2021) were concentrates of filtrates, while the fifth (February 2021) was a concentrate of FTS (Table 1).

**Table 1:** Concentrates analyzed in this study and virus detection by nested PCR. ‘+’ indicates virus was detected

| Month | Conc ID | Filtrate |  |  | Filter-Trapped-Solid |  |  |  | Total |
| --- | --- | --- | --- | --- | --- | --- | --- | --- | --- |
|  |  | EV | CanPV | Ad41 | Conc ID | EV | CanPV | Ad41 |  |
| Oct 2019 | 1 | + |  | + | 13 | + | + | + | + |
| Nov 2019 | 2 | + |  | + | 14 | + | + | + | + |
| Dec 2019 | 3 | + | + | + | 15 | + | + | + | + |
| Jan 2020 | 4 | + | + | + | 16 | + | + | + | + |
| Feb 2020 | 5 |  |  | + | 17 | + |  | + | + |
| Mar 2020 | 6 | + | + | + | 18 | + | + | + | + |
|  | Season 1 | **5/6** | **3/6** | **6/6** |  | **6/6** | **5/6** | **6/6** | **6/6** |
| Oct 2020 | 7 |  |  |  | 19 | + |  |  | + |
| Nov 2020 | 8 |  |  |  | 20 | + | + |  | + |
| Dec 2020 | 9 | + | + |  | 21 | + | + |  | + |
| Jan 2021 | 10 | + | + |  | 22 | + | + |  | + |
| Feb 2021 | 11 | + |  | + | 23 |  |  |  | + |
| Mar 2021 | 12 |  | + | + | 24 | + |  |  | + |
|  | Season 2 | **3/6** | **3/6** | **2/6** |  | **5/6** | **3/6** | **0/6** | **6/6** |
|  | Total | **8/12** | **6/12** | **8/12** |  | **11/12** | **8/12** | **6/12** | **12/12** |

#### EV typing

Post-trimming, 36,565,524 Illumina raw reads (Table S2) were determined for the 19 samples, and 81 EV variants (assembled from 64.72% of the trimmed reads) were recovered (Table 2). The 81 variants belonged to 31 EV types and five species (Table 2). Ninety-four percent (29/31) and 96% (78/81) of the EV types and variants detected, respectively, belonged to EV-A, B and C. EV-B (12 types and 17 variants) and EV-C (10 types and 42 variants) had the largest number of types and variants detected, respectively (Table 2).

**Table 2:** EV variants detected per Species 2019/2020 and 2020/2021.

| S/N | Species | Types | Total types | Total variants |
| --- | --- | --- | --- | --- |
| 1 | EVA | CVA2(3), CVA5(4), CVA6(3), CVA10(2), CVA16(2), EVA76(4), EVA90(1) | 7 | 19 |
| 2 | EVB | CVA9(1), CVB2(3), CVB3(1), CVB4(2), CVB5(1), E3(1), E6(1), E9(1), E11(1), E14(2), E18(1), E30(2) | 12 | 17 |
| 3 | EVC | CVA1(6), CVA11(7), CVA13(5), CVA17(1), CVA19(11), CVA20(1), CVA21(3), CVA22(4), CVA24(1), EVC99(3) | 10 | 42 |
| 4 | RVB | B79 | 1 | 1 |
| 5 | RVC | C36 (2) | 1 | 2 |
| **Total** |  |  | 31 | 81 |

#### EV diversity by fraction

We found that all months in both seasons were positive for EVs, but not all fractions. For season 1, 65 EV variants (belonging to 28 EV types and four species) were determined, 46 from FTS (from 24 EV types and five species) and 19 from filtrates (from 12 EV types and three species) (Figures 2, 3, 4a and 4b). Eight EV types belonging to three species were determined from both fractions (Figure 4c). However, 16 and four were uniquely detected in FTS and filtrates, respectively (Figure 4c). EV types from five species (EV-A, EV-B, EV-C, RV-B and RV-C) were recovered from FTS, while only three species (EV-A, EV-B and EV-C) were determined from filtrates (Figure 4c). Also, in four of the six months in season 1, the same EV types and variants were recovered from both concentrates. However, EV type and diversity in FTS were not consistent with that in the filtrate and vice-versa (Figure 2).

**Figure 2:**
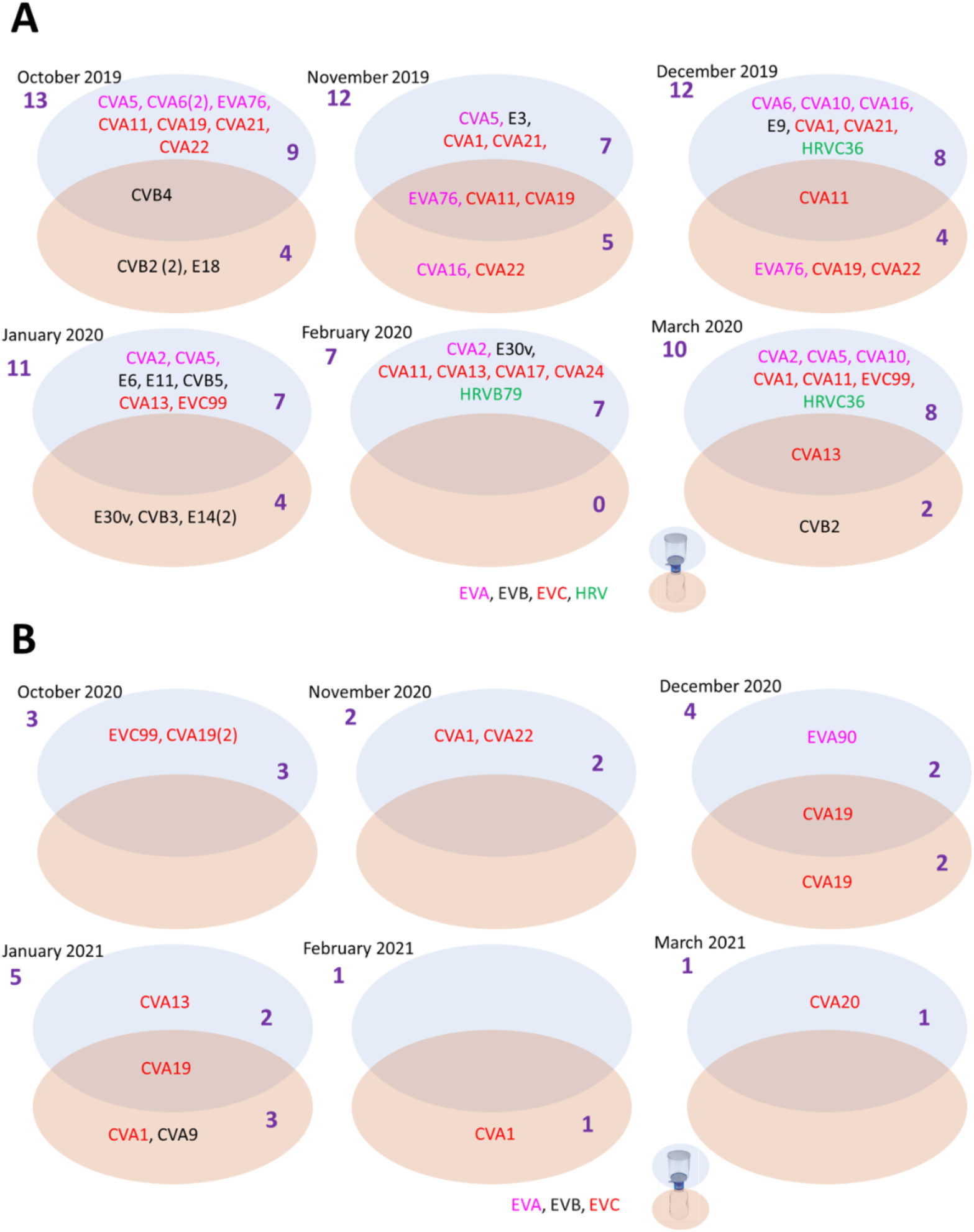
EV variants detected in different fractions of WW. A) October 2019 to March 2020. B) October 2020 to March 2021. Variants are colored by species according to the legend on the bottom. For each month, EV types/variants detected in FTS or filtrate are clustered in blue or beige ovals, respectively. Those detected in both are in the overlapping region of both ovals.

**Figure 3:**
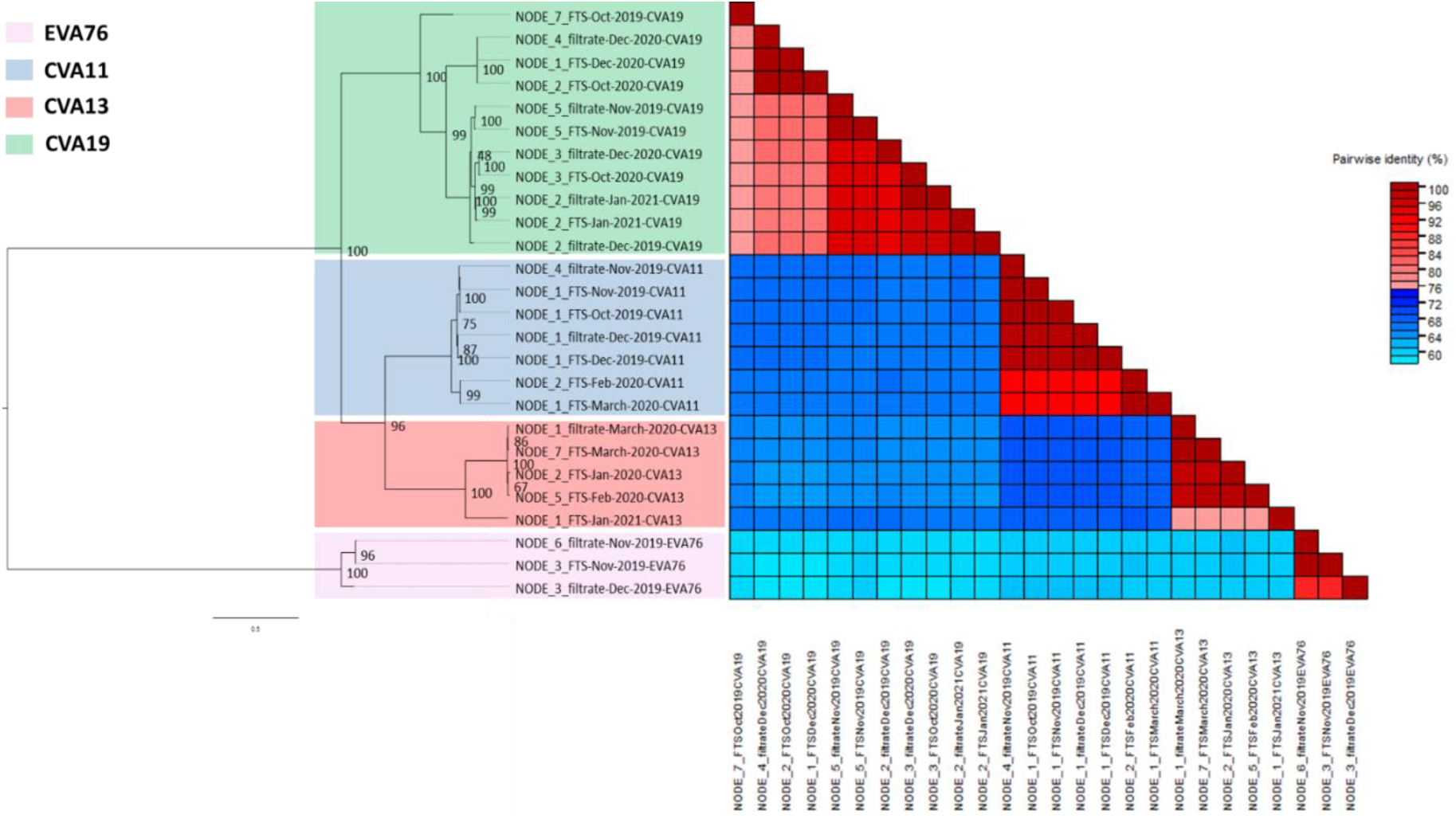
Genetic characterization of EV variants (and types) that were present in both fractions in Figure 2. The ML tree was inferred using IQ-Tree 1.6.12. with best substitution model (TIM2+F+G4) selected using ModelFinder. Pairwise similarity analysis was done using SDT with both cut-offs in 3-color mode set at 75% (i.e., 25% divergence).

**Figure 4:**
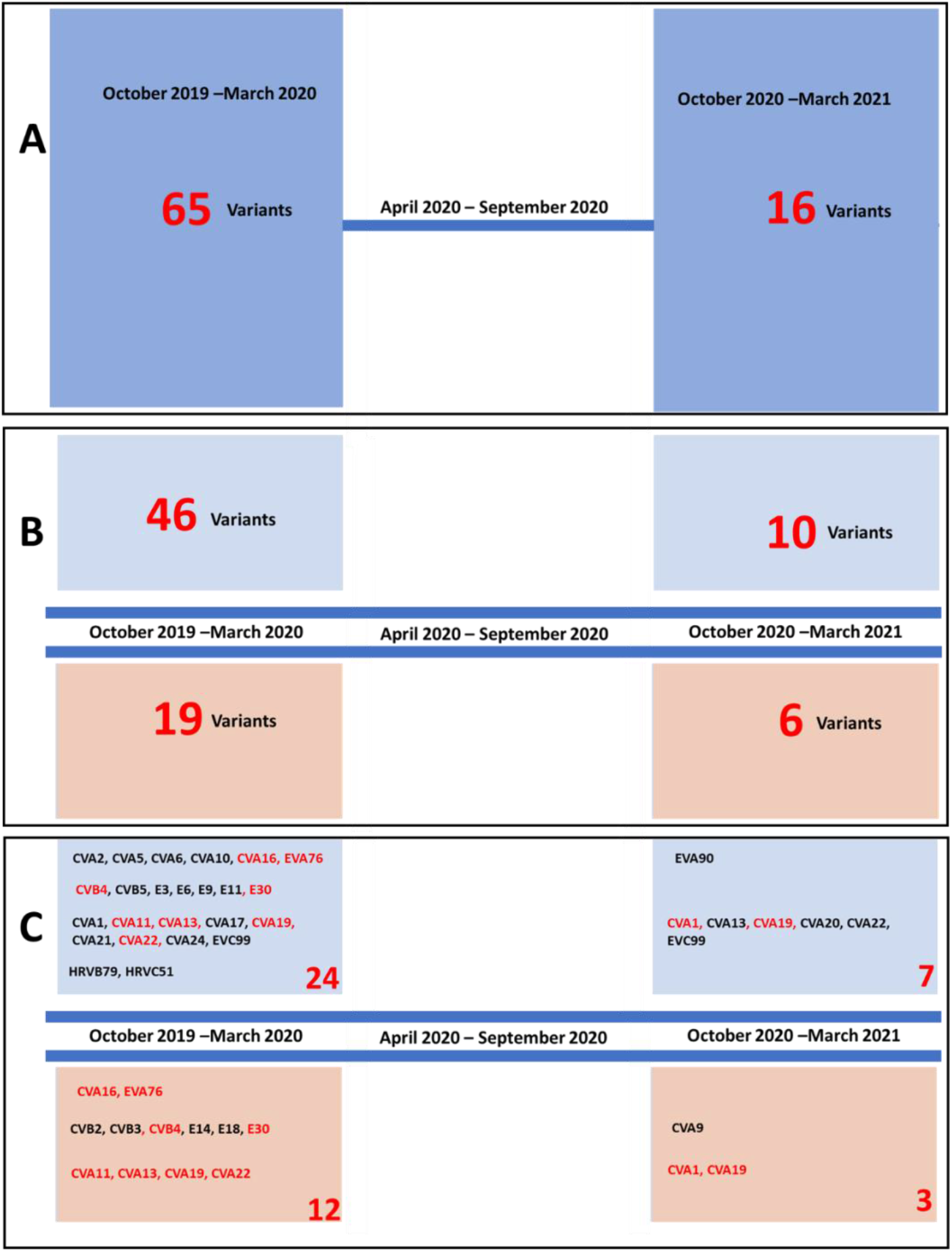
EV diversity by season. A) total number of variants detected in each season. B) Total number of variants detected in each fraction during each season. C) The different EV types detected in each fraction during each season. EV types detected in both fractions are in red font. In B and C, EV types/variants detected in FTS or filtrate are clustered in blue or beige rectangles, respectively.

For Season 2 (S2), EVs were detectable in all FTS samples except for that from February 2021, but only filtrates from December 2020, January 2021 and February 2021 yielded EVs. For season 2, 16 EV variants (from eight EV types and three species) were determined, 10 from FTS (from 7 EV types and two species) and six from filtrates (3 EV types and 2 species). Two EV types were determined from both fractions, five and one were uniquely detected in FTS and filtrates, respectively. EV types from two species (EV-A and EV-C) were recovered from FTS. The same was observed for filtrates but from EV-B and EV-C. Also, in half of the months, some EV types and variants were recovered from both concentrates. However, EV type and diversity in FTS were not consistent with that in filtrate and vice-versa (Figure 2).

There was a 3.9x drop in EV variants detected between seasons 1 and 2. Precisely, 63 and 16 EV variants were detected in seasons 1 and 2, respectively. There was a 3.5x drop in EV types detected between seasons 1 and 2. Exactly, 28 and 8 EV types were detected in seasons 1 and 2, respectively. It is important to mention that only 5 of the 28 EV types detected in season 1 were detected in season 2. Three (CVA9, CVA20 and EVA90) of the EV types detected in season 2 were not detected in season 1 (Figure 4).

### CANINE PICORNAVIRUSES

We detected CanPVs in 58.3% (14/24) of the samples suggesting CanPVs were present (Table 1). Both FTS and filtrate of February 2020 and 2021, and October 2020 (six samples in all) were negative for CanPV. The four remaining negative samples were filtrates from October 2019, November 2019, November 2020, and FTS from March 2020. Post-trimming, we had 13,824,046 Illumina raw reads (Table S3) for the 14 samples from which 27 CanPV contigs (assembled from 59.91% of the trimmed reads) were recovered (Figures 5, 6 and 7).

**Figure 5:**
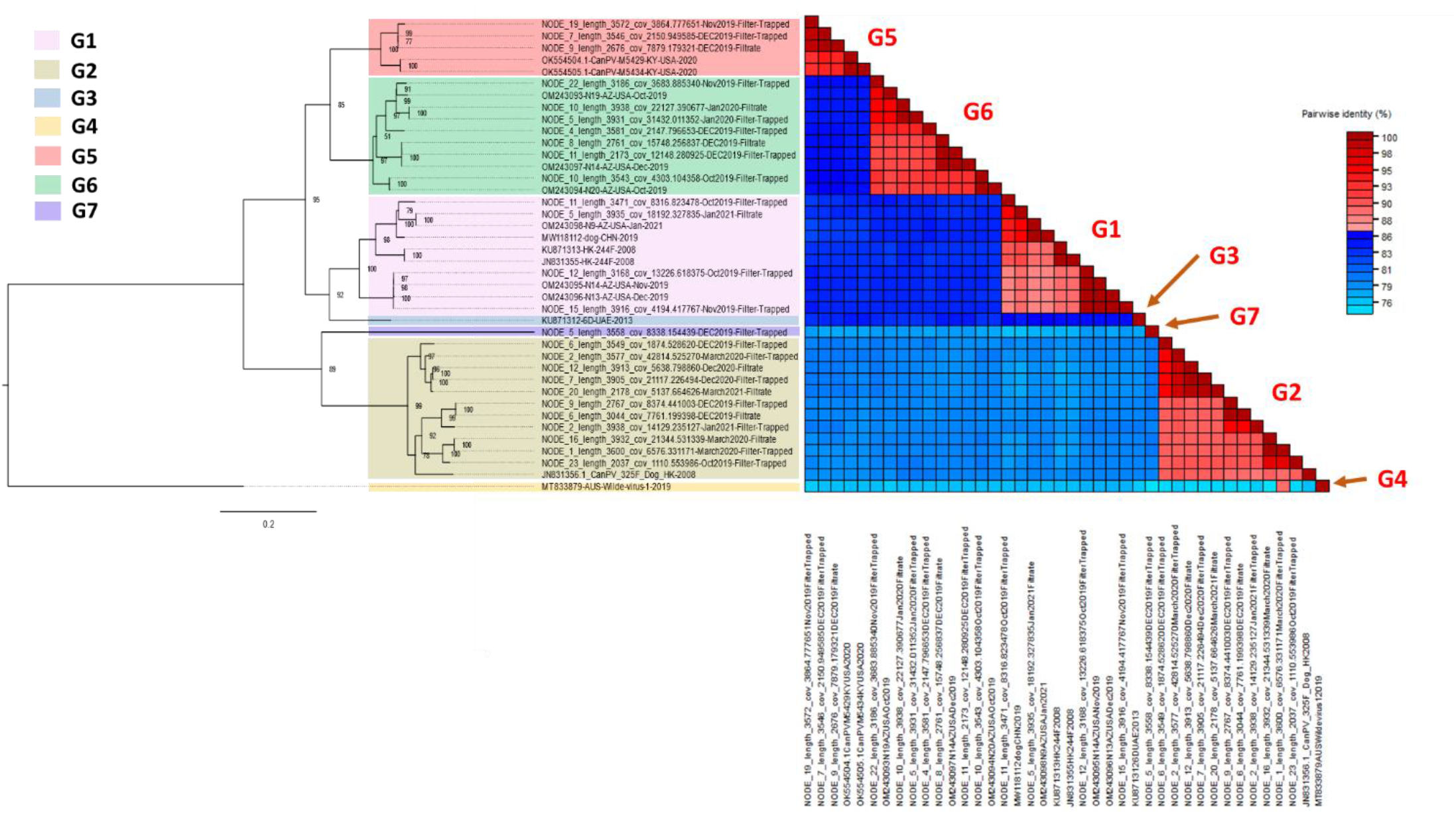
Genetic characterization of CanPV variants. Maximum-Likelihood (ML) tree and pairwise similarity analysis of CanPV variants present in GenBank and those recovered in this study. The ML tree was inferred using IQ-Tree 1.6.12. with the best substitution model (TIM2+F+I+G4) selected using ModelFinder. Pairwise similarity analysis was done using SDT with both cut-offs in 3-color mode set at 86% (i.e., 14% divergence).

**Figure 6:**
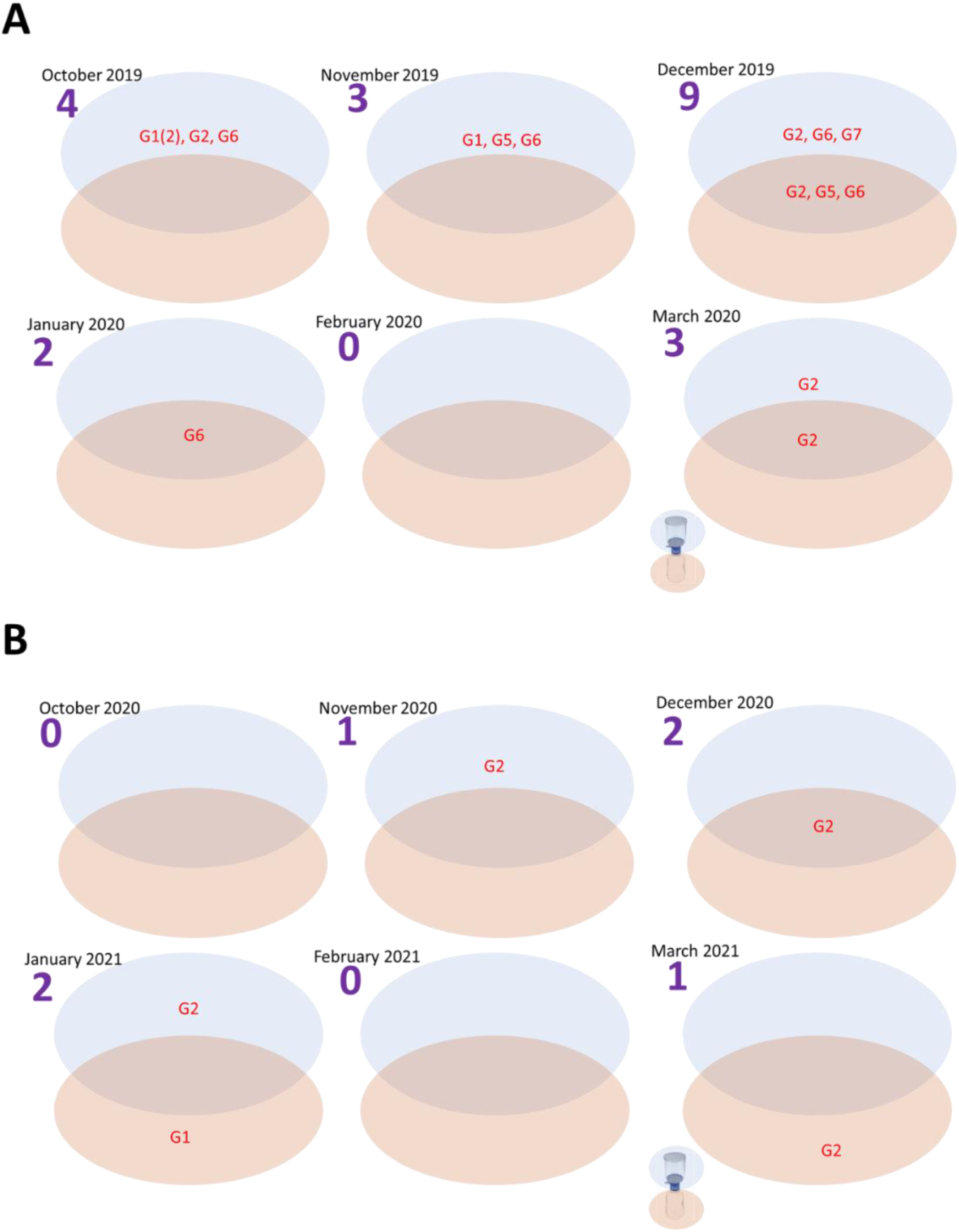
CanPV variants detected in different fractions of WW. A) October 2019 to March 2020. B) October 2020 to March 2021. For each month, CanPV types/variants detected in FTS or filtrate are clustered in blue or beige ovals, respectively. Those detected in both are in the overlapping region of both ovals.

**Figure 7:**
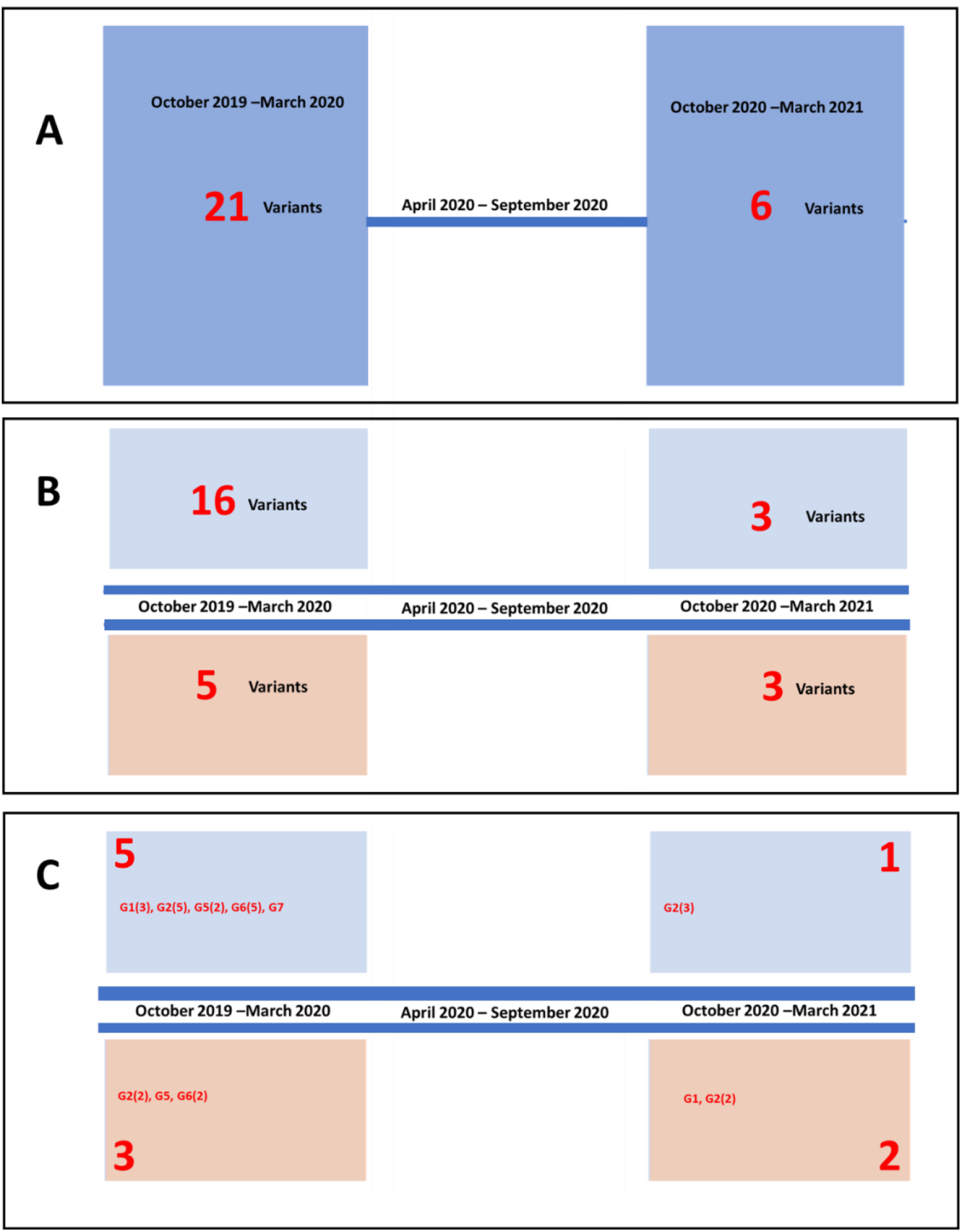
CanPV diversity by season. A) Total number of variants detected in each season. B) Total number of variants detected in each fraction during each season. C) The different genotypes detected in each fraction during each season. For B and C, CanPV types/variants detected in FTS or filtrate are clustered in blue or beige rectangles, respectively.

#### CanPV typing

Using phylogenetic and pairwise identity analysis, we found that the CanPV sequences generated in this study and those publicly available in GenBank form seven (7) distinct clusters with strong bootstrap support (Figure 5). Divergence within each of the seven clusters was less than 14% (Figure 5). Though we currently do not know what this clustering pattern represents with respect to the biology of CanPV, it provides a useful tool for classification, which we utilize henceforth for genotyping CanPV variants in this study. We assign these genotypes historically. Hence, genotypes are numbered based on the year of detection of the oldest variant within the cluster.

Our data, therefore, show that genotypes G3 and G4 have only been described in the United Arab Emirates (UAE) and Australia, respectively to date^21,22^. Genotype G1 has been previously described in Hong Kong and China and G2 in Hong Kong^20,21,23^. Genotypes G5 and G6 had been previously described in the USA^24,25^. This study shows that genotypes G1, G2, G5, G6 and G7 are present and likely circulating in the USA. Based on publicly available sequence data in GenBank, this is the first description of genotype G7 (Table 3).

**Table 3:**
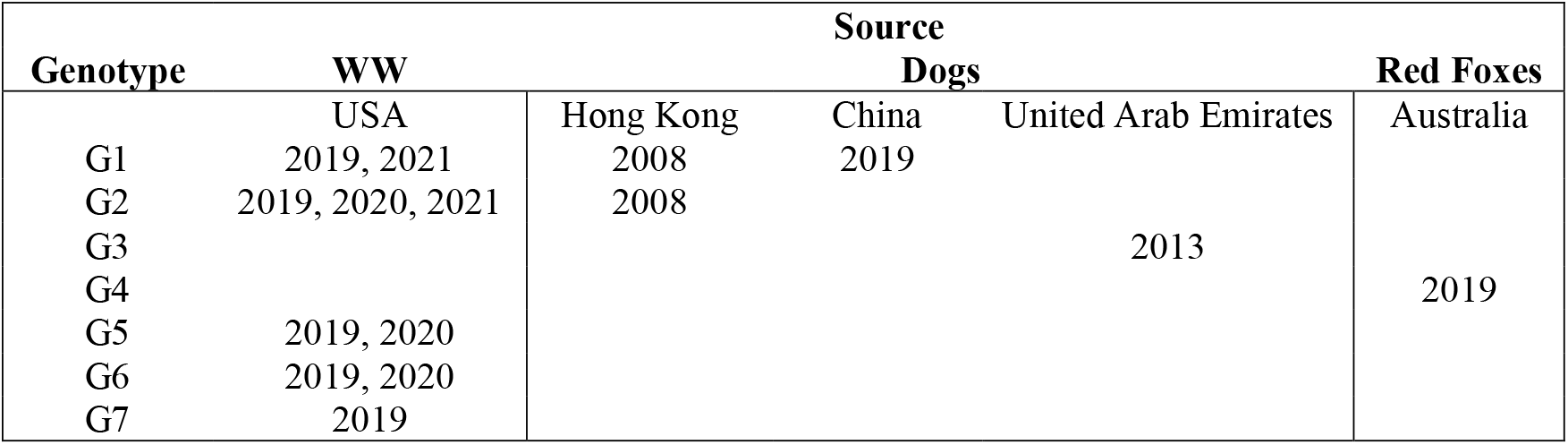
Global distribution of CanPV types described to date based on sequence data publicly available in GenBank as of July 2022. Note that members of each genotype have divergence below 14%. Please see Figure 5.

#### CanPV diversity by fraction

For season 1, 21 CanPV variants (belonging to 5 genotypes) were detected, 16 from FTS (from 5 genotypes) and five from filtrates (from three genotypes). All three CanPV genotypes detected from filtrates were also recovered from FTS. However, two CanPV genotypes (G1 and G7) were detected from only FTS while none was uniquely detected from filtrates. In summary, for season 1, CanPV genotype and variant diversity detected in FTS was consistent with what was found in filtrate, but genotype and variant diversity detected in the filtrate was not consistent with what was found in FTS (Figures 6 and 7).

For season 2, 6 CanPV variants (belonging to 2 genotypes) were detected, three each from FTS and filtrate. Genotype G2 was detected in both FTS and filtrates. However, genotype G1 was also detected in filtrates but not in FTS. In December 2020 the same CanPV variant was detected in both FTS and filtrates. For the other months (except for October 2020 and February 2021 where both fractions were negative), different CanPV variants were found in both fractions. In summary, for season 2, CanPV type and diversity detected in FTS was not consistent with what was found in the filtrate and vice-versa (Figures 6 and 7).

A 3.5x decrease in CanPV variants was detected between seasons 1 and 2. Twenty-one and six CanPV variants were detected in seasons 1 and 2, respectively. Only two (G1 and G2) of the five CanPV types detected in season 1 were detected in season 2. The remaining three CanPV genotypes (G5, G6 and G7) were not detected in season 2 (Figure 7).

### HUMAN Adenovirus 41

#### Fractions positive for Ad41

We detected human Ad41 in 58% (14/24) of the samples suggesting the presence of human Ad41 (Table 1). Of the 10 negative samples, four (October-December 2020, January 2021) were concentrates of filtrates, while six (October-December 2020, January-March 2021) were concentrates of FTS (Table 1).

All season one samples (irrespective of fraction and month) were positive for the Ad41 nested PCR assay (Table 1). For season 2, only filtrates for February and March 2021 were positive for the second-round (fiber) assay (Table 1), suggesting there must have been amplification in the first round Ad41 CG assay.

#### Analysis of Illumina sequencing data

In all, 30,277,612 Illumina raw reads resulted from the 14 samples. 13,797,291 (45.57%) of these raw reads mapped to the reference (human adenovirus 41 isolate MU22/patient E; MW567966) Ad41 complete genome (Figure S1). To determine human adenovirus 41 (MW567966) as the reference sequence for template-guided assembly, the raw reads were first *de novo* assembled, and contigs recovered were used as a query via a BLASTn search of the GenBank database. Ad41 (MW567966) was the most similar complete genome in GenBank. It was consequently selected and used as template for template-guided assembly.

Template-guided assembly showed that assay 4 (which covers the genomic region containing the penton protein complete coding sequence) of the complete genome assay failed in all cases. Hence, as opposed to amplifying about 97.5% of the genome (in accordance with assay design), assay 4 failure resulted in the CG assay yielding about 87.5% of Ad41 genome (Figure S1).

The penton, hexon and fiber genes are usually the most dynamic Ad41 capsid proteins. However, since assay 4 failed (and consequently, we could not recover the penton gene for all our samples) we investigated the variant profile of hexon and the two (small and large) fiber genes. Variant analysis of the genes showed seven, one and three unique profiles for the hexon, small fiber and long fiber genes, respectively (Tables 4, 5, 6). The variability detected in the hexon gene region clustered in the hypervariable region (HVR) (Table 4).

**Table 4:**
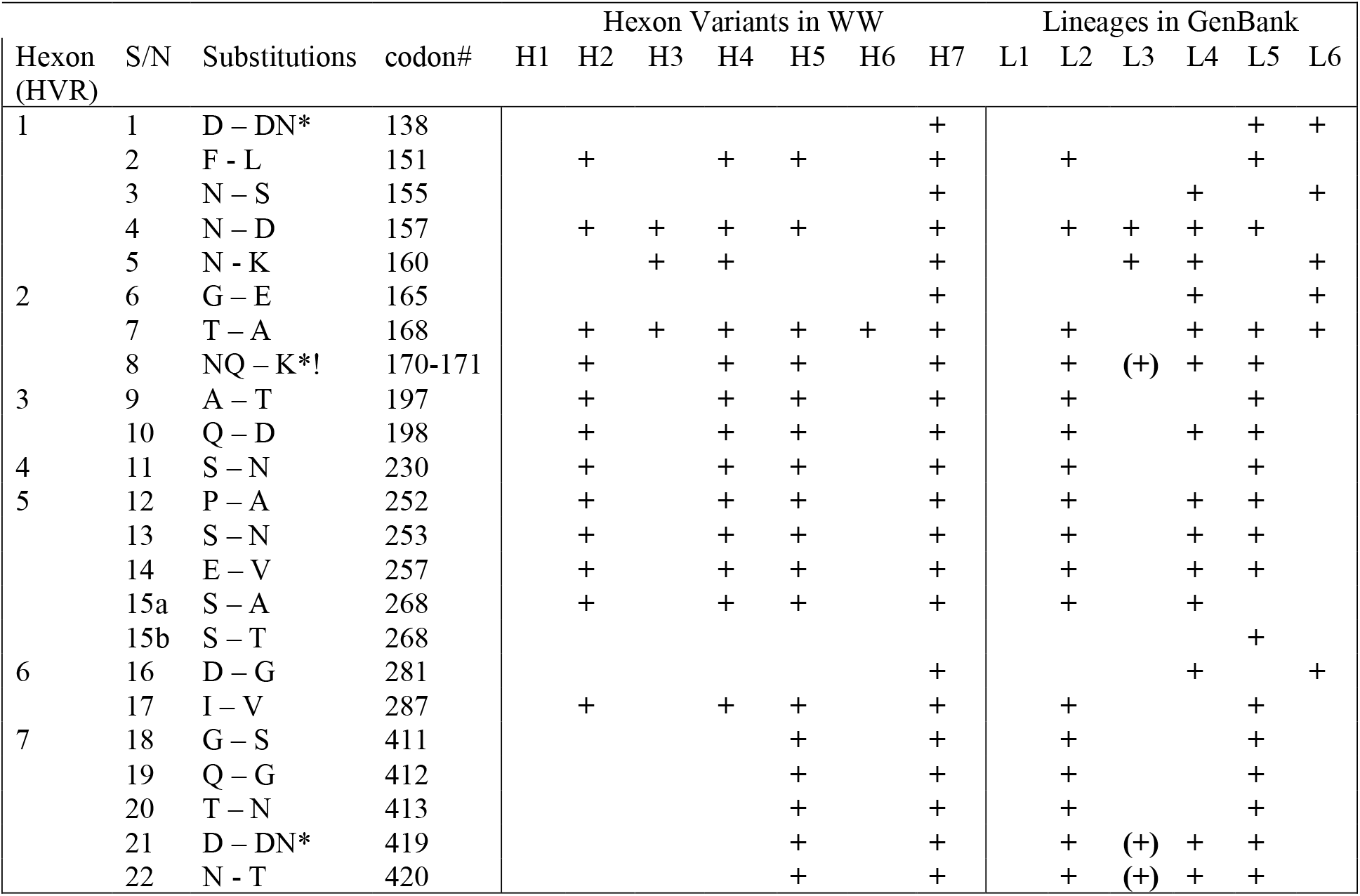
Amino acid substitutions in hexon gene hypervariable region (HVR) detected by variant analysis of mapped reads in this study and their presence in Ad41 lineages in GenBank. Codon numbering is relative to the hexon gene of MW567966 (L1). Note that L1 has the same sequence as H1. Also, *=Insertion, *! = Insertion and deletion, + = present in all members, (+) = present in all some members. Please note that all variant sites called had >1000x coverage.

**Table 5:** Amino acid substitutions and deletions detected by variant analysis of mapped reads in long-fiber. Codon numbering is relative to long fiber gene of MW567966 (L1). Note that L1 has the same sequence as the reference. Hence, will have no substitutions and is consequently not listed in this table.

| **S/N** | **Sub** | **codon#** | **L2** | **L3** |
| --- | --- | --- | --- | --- |
| 1 | F – V | 251 | + |  |
| 2 | 15aa deletion | 263-277 |  | + |

**Table 6:** Long fiber gene variant profile of WW concentrates from October 2019 to March 2020.

| **Month-Year** | **FTS** | **Filtrate** |
| --- | --- | --- |
| Oct-19 | L1 | L1 + L2 |
| Nov-19 | L1 + L2 | L1 + L2 |
| Dec-19 | L1 | L1 |
| Jan-20 | L1 + L3 | L1 |
| Feb-20 | L1 | L1 |
| Mar-20 | L1 | L1 |

For the long fiber gene, the F251V substitution was the only amino acid substitution detected by variant analysis (Table 5) and when present was between 38% and 55% of mapped reads in all samples. Hence, this suggests that the F251 was equally present in the population. The long fiber phylogenetic tree showed that variants with F251V substitution formed a distinct cluster (Figure 10). Variant analysis also showed the presence of a variant with a 45nt (15aa) deletion in the fiber gene in January 2020. However, it was only found in FTS and not the filtrate (Table 6). For the small fiber gene, the L362F substitution was the only amino acid substitution detected by variant analysis and was present in more than 99% of raw reads in all samples.

The seven variant profiles of hexon genes recovered from the concentrates analyzed in this study were very dynamic, containing between one and 22 amino acid substitutions that formed seven distinct patterns (H1-H7) (Table 4). Only in the January 2020 sample did the hexon gene variant profile of FTS match the filtrate. In all other months in season 1 they were different (Figure 8).

**Figure 8:**
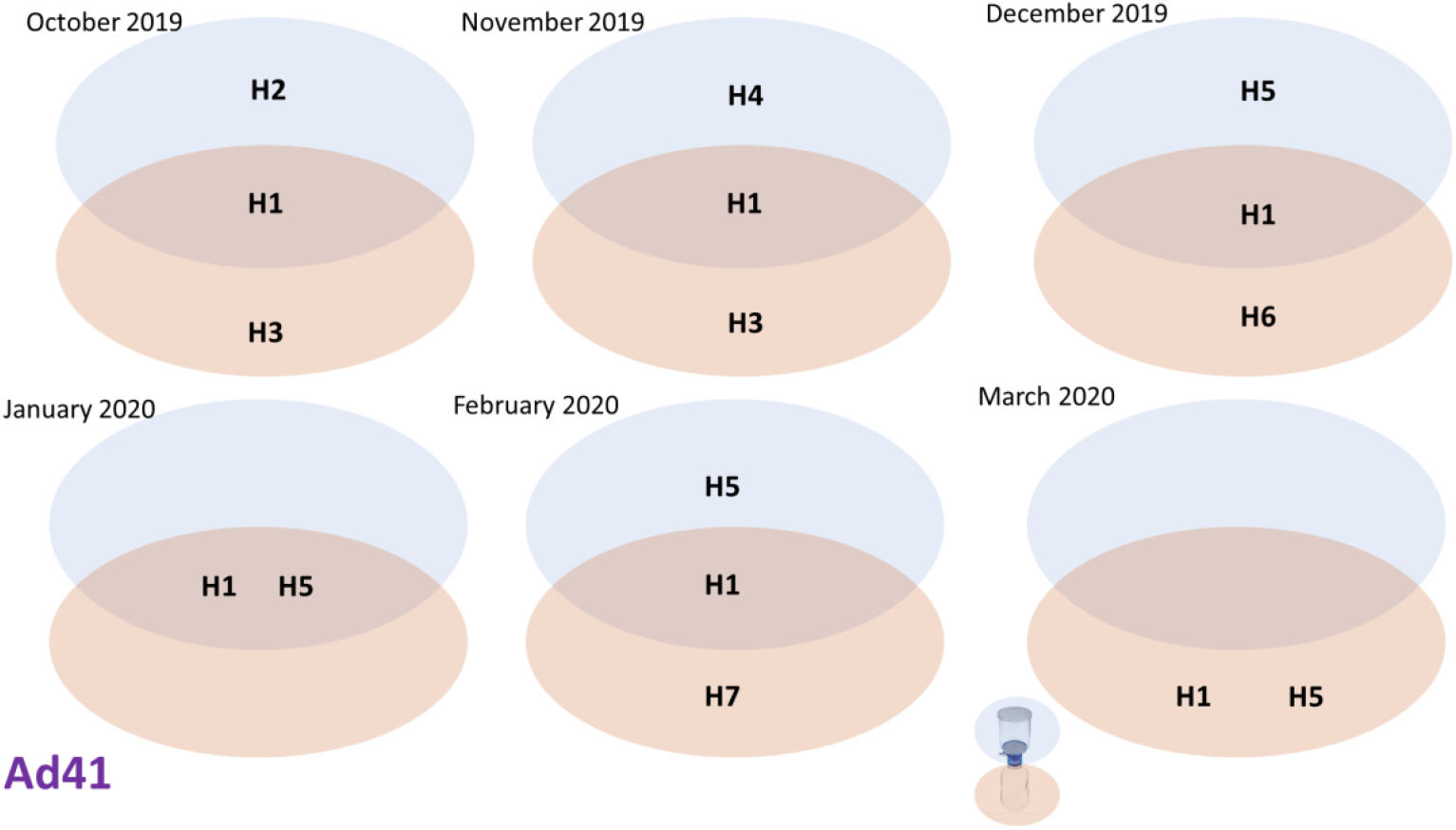
Hexon gene variants identified in different fractions of WW from October 2019 to March 2020. Please note that FTS from March 2020 had Ad41 present based on 2^nd^ round PCR result (Figure S2c). However, only assay 7 (which captures short and long fiber coding region but not hexon) worked in the first-round assay (Figure S1). For each month, Ad41 types/variants detected in FTS or filtrate are clustered in blue or beige ovals, respectively. Those detected in both are in the overlapping region of both ovals.

#### Analysis of GenBank deposited data

To understand how the variant profiles found in WW in this study track with those present in variants publicly available in GenBank, we collected hexon gene complete coding sequences of the top 100 variants recovered from a GenBank search using MW567966 as the query. Precisely, 59 of the variants were extracted from complete genomes while the remaining 41 were not part of complete genomes but had the complete hexon coding sequence publicly available in GenBank. Phylogenetic and pairwise similarity analyses suggest the Ad41 hexon gene clustered into six (L1-L6) lineages (Figure 9). Intra-lineage divergence was less than 0.4% (Figure 9) and each lineage had a unique combination of amino acid substitutions (Table 4).

**Figure 9:**
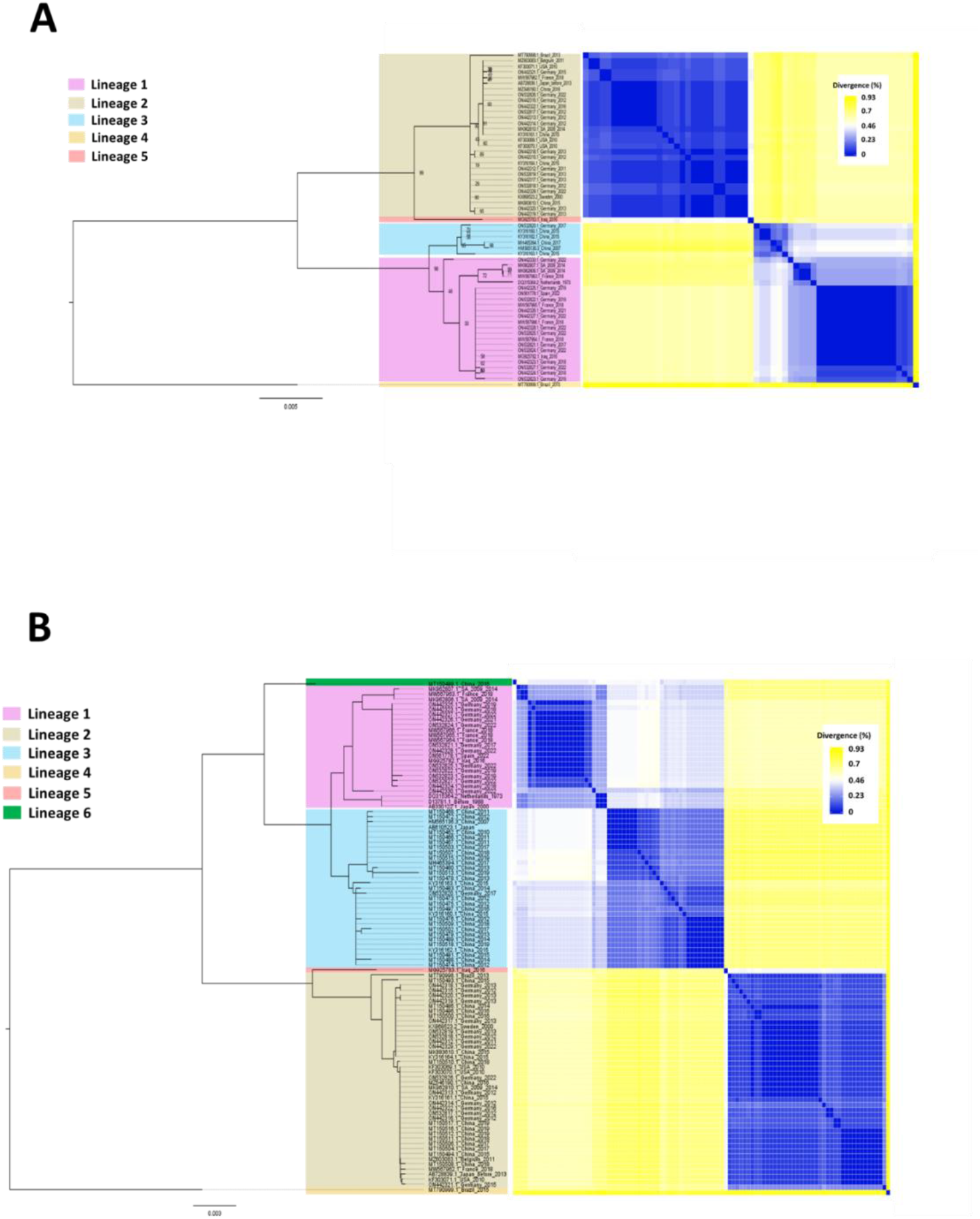
Genetic characterization and similarity of Ad41 Hexon gene variants. Maximum-Likelihood tree and pairwise divergence analysis of Ad41 A) Hexon genes extracted from complete genomes in GenBank and B) top 100 hits of a BLASTn search of the GenBank database using MW567966 as the query sequence.

When we compared the amino acid variation profiles of hexon genes found in WW in this study with those from complete genomes in GenBank, we found that H1= L1, H5 = L2, H3 is a subset of L3 and H7 = L5 + L6. We also found that the amino acid variation profiles H2, H4, and H6 were not represented in lineages 1-6 (Table 4).

A comparison of phylogenetic trees generated independently for hexon, short fiber and long fiber from the 59 complete genomes showed phylogeny violation suggesting that recombination might be occurring between the hexon gene and the fiber genes (Figure 10), which are over 8kb apart. Particularly striking were the phylogenetic relationships of MG925783 (Iraq 2016), MK962807 (South-Africa 2009-2014), MK962808 (South-Africa 2009-2014), MW567963 (France 2018), and ON442330 (Germany 2022). In the hexon gene tree, MG925783 (Iraq 2016) is the only member of lineage 5 while the other four variants belong to two subclusters of lineage 1 (Figure 10). However, in the long fiber tree all five sequences form a distinct cluster (Figure 10). Similarly, the five sequences form a distinct cluster in the tree, made using concatenated (hexon-small fiber-long fiber) sequences (Figure 10). Further investigation showed that all five variants share a related long-fiber gene (with the characteristic 45 nucleotide [and consequently 15aa] deletion that results in the loss of one turn of the fiber shaft (Figure 11)). Interestingly, ON442330 does not cluster with the other four (MG925783, MK962807, MK962808, and MW567963) in the small fiber gene tree (Figure 10).

**Figure 10:**
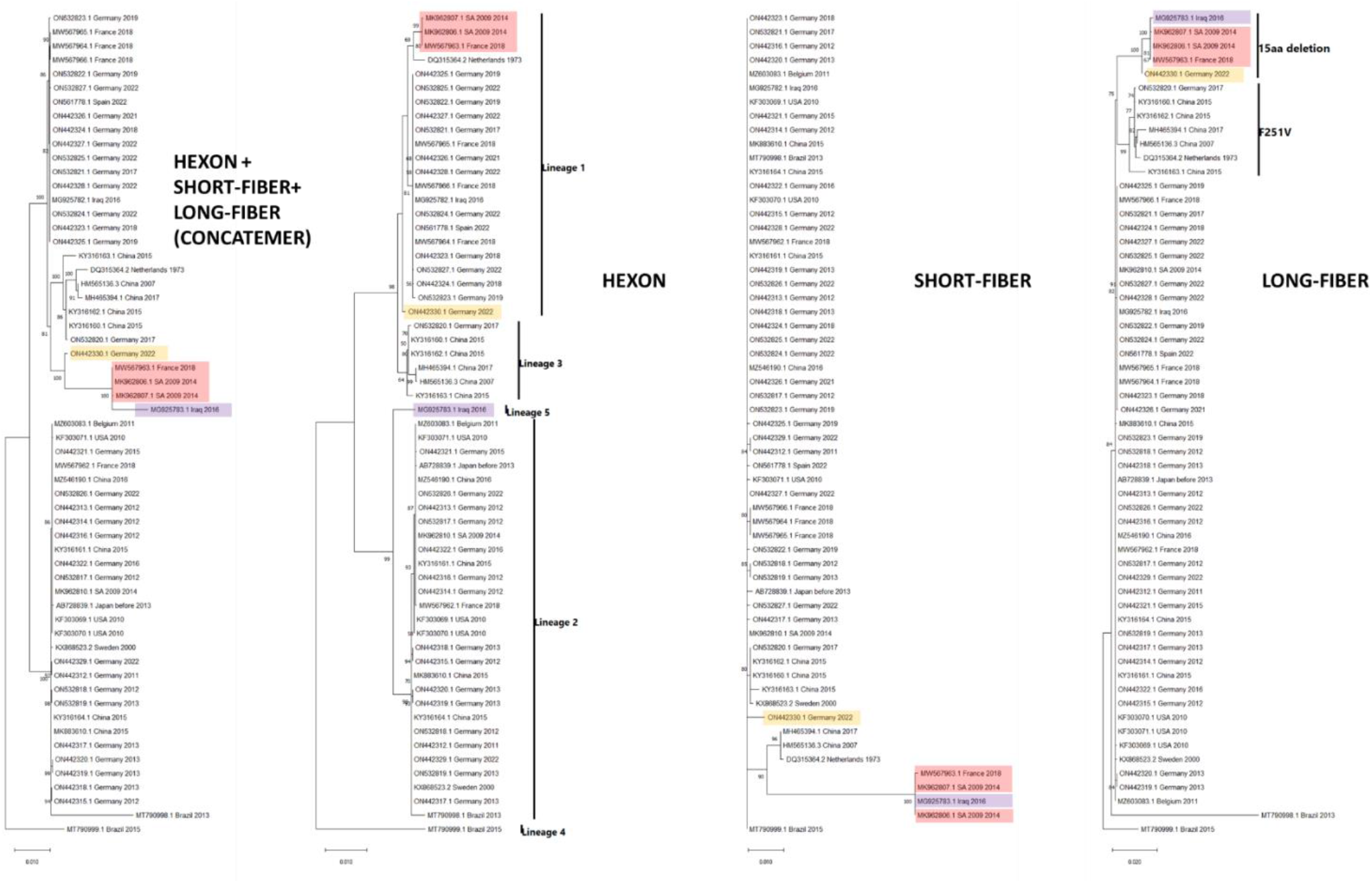
Maximum-Likelihood tree of Ad 41 variants. The first tree on the left was inferred using concatenated hexon, small fiber and long fiber protein coding gene sequences extracted from 59 Ad41 complete genomes present in GenBank. The tree was inferred using IQ-Tree 1.6.12. with the best substitution model selected for each of the partitions using ModelFinder. The remaining trees are of hexon (K3P+I substitution model), small fiber (HKY+F substitution model) and long fiber (HKY+F substitution model) protein coding gene sequences extracted from 59 Ad41 complete genomes present in GenBank. These trees were also inferred using IQ-Tree 1.6.12. with the best substitution model selected using ModelFinder. Bootstrap values are shown if greater than 50%. Sequences of interest are color-coded to highlight phylogeny violations between the different trees.

**Figure 11:**
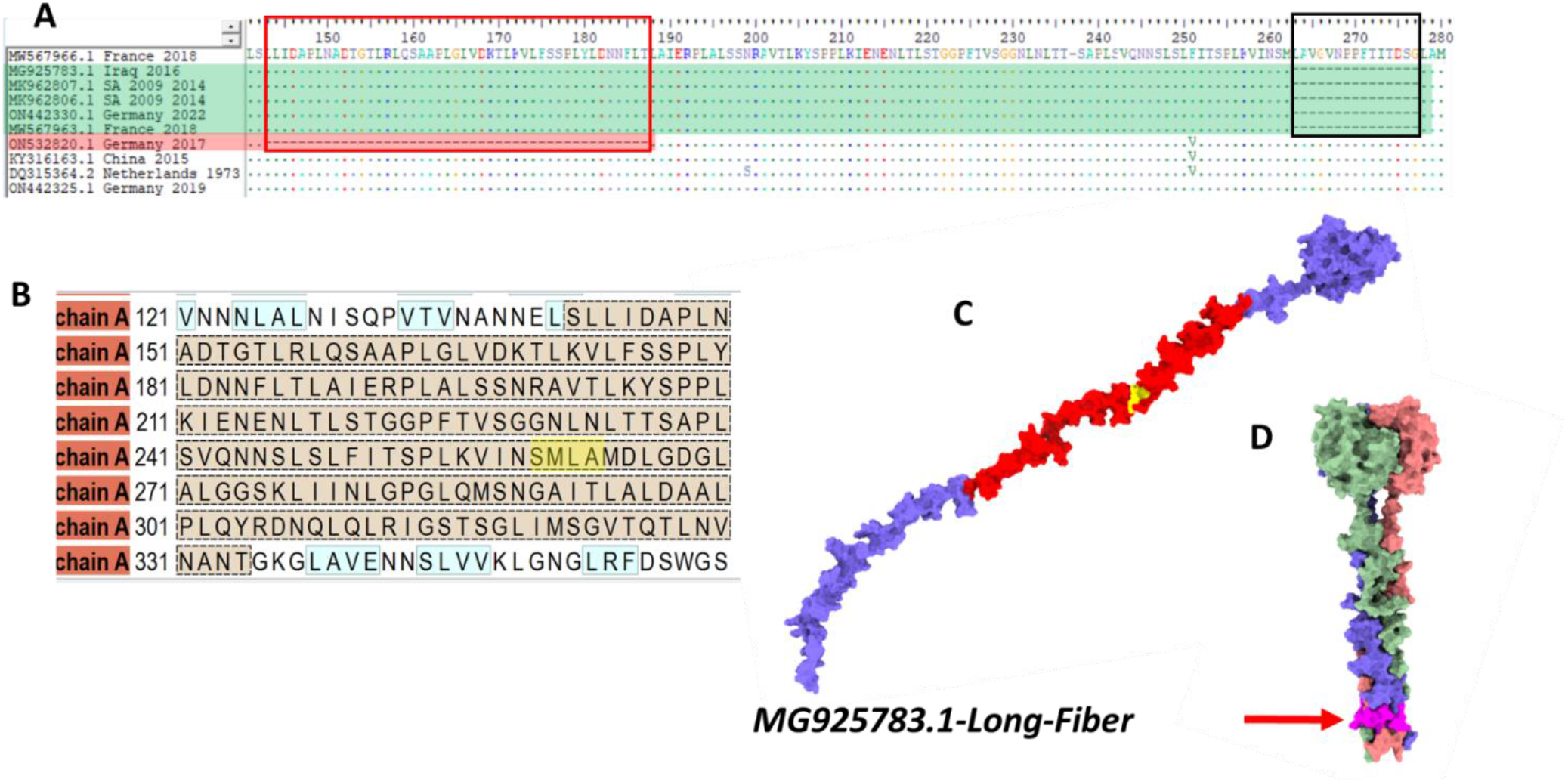
Amino acid (aa) analysis of the long fiber of Ad41. A) alignment of a subset of sequences (amino acid 141-280) described in the phylogenetic trees (Figure 10). MG925783, MK962807, MK962808, MW567963 and ON442330 are highlighted in green. Please note the 15aa deletion (LAVGVNPPFTITDSG [black rectangular highlight]) flanked on the N and C termini by SM and LA, respectively. Also note the 45aa deletion (red rectangular highlight) in ON532820 (red highlight). B) A portion of the aa sequence of MG925783 highlighting in brown and yellow the region amplified by the 2^nd^ PCR assay (Figure 1) and amino acids SM and LA (flanking the 15 aa deletion), respectively. Note that the first 45aa of the region amplified by the Ad41 2^nd^ round PCR corresponds to the 45aa deletion in ON532820 suggesting that the assay will miss sequences with this deletion. C) ColabFold predicted structure of MG925783 long fiber monomer. Highlighted in red and yellow are the region amplified by the 2^nd^ round PCR assay and the aa (SM and LA) flanking the 15aa deletion, respectively. D) ColabFold predicted trimeric structure of the c-terminal 200aa of long fiber consensus sequence recovered from October 2019 in this study. The portion of the long fiber shaft corresponding to the 15aa deletion is highlighted in pink and pointed to using a red arrow.

To understand how the Ad41 variants recently recovered from children with hepatitis-of-unknown origin fit into this schema, the published^27^ hexon genes (ON565007-ON565011) were aligned with representatives of the six lineages. Our data shows that they belong to lineages 1, 2 and 6 with both genomes recovered from children with acute liver failure belonging to lineage 6 (Figure 12).

**Figure 12:**
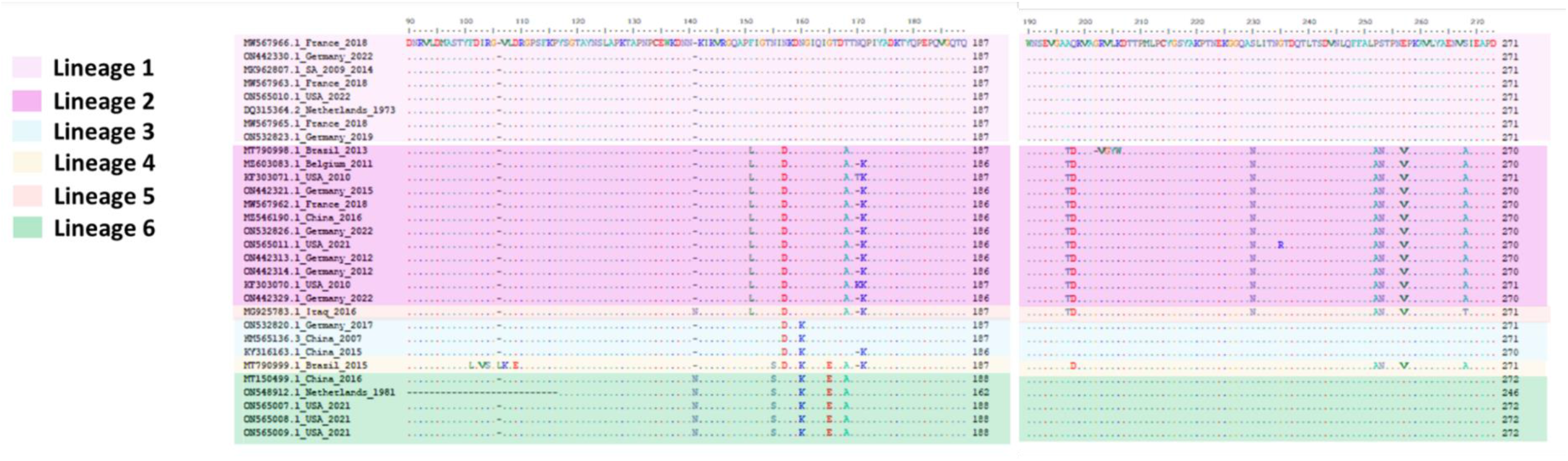
A subset of the unique amino acid substitution signatures of Ad41 hexon gene protein sequence from representatives of lineages 1-6 (residues 90 – 271; MW567966 numbering, and publicly available sequences of variants recovered from children with hepatitis of unknown origin^27^.

## Discussion

### FTS vs filtrate

We studied (using three different viruses) whether size fractionation of wastewater samples prior to ultrafiltration impacts our perception of virus presence and diversity in wastewater samples. Our results consistently show that it does reveal a difference (Figures 3, 4, 6, 7 and 8). Firstly, at least one of the three virus types screened for in this study was detected in concentrates from each of the months sampled. However, not all partitions had virus. For example, while EV and CanPV were detected in FTS from November 2020, no virus was detected in the corresponding filtrate. On the other hand, while EV and Ad41 were detected in filtrate from February 2021, no virus was detected in FTS from the same month (Table 1). Hence, with respect to virus presence or absence in the WW sample, both (FTS and filtrate) partitions seem not to be consistent with each other (Table 1).

Previous studies^7,8,9,10,11^ investigating the impact of the clarification process have assumed that virus presence and diversity are one and the same. Hence, they did not examine how the clarification process impacts our perception of virus diversity in both fractions. In this study, our results showed that even in cases where virus was detected in both partitions, virus diversity captured either in FTS or filtrate seem not to be consistent with each other. For example, while 12 different EV types were detected in December 2019, seven and three were uniquely detected in the FTS and filtrate, respectively. Only one (CVA11; same type and variant) was detected in both (Figures 2 and 3). Furthermore, when summed over the course of either or both seasons, our data showed that virus types could be present in a population and WW clarification by size fractionation could determine whether it is detected over the course of a six-month period (at least based on this study). For example, in season one, 16 EV types were present in the population sampled that were not detected in the filtrate. Similarly, four EV types were present in the filtrate that were not detected in the FTS (Figure 4). A similar phenomenon was also observed for CanPV and Ad41 (Figures 6, 7 and 8). Such delay can undermine WBE’s function as an early warning system by delaying our ability to detect variants circulating in the population. Our findings therefore show that while (in an effort not to clog the pores in centrifugal filters) sample clarification (by size fractionation) is essential prior ultrafiltration, it might be creating an artificial representation of viral diversity in the resultant sample. We therefore recommend virus recovery from both partitions (FTS and filtrate) for an accurate representation of virus presence and diversity in WW samples.

The abundance of large, particulate fecal matter in the primary clarification chamber of wastewater treatment plants (WWTPs) shows that not all fecal matter in WW completely dissolves and release trapped virus particles into suspension. Hence, the possibility exists that some of the viruses recovered in FTS might have been those trapped in undissolved fecal matter or even adsorbed to solids in WW. Additionally, it has been shown that enteric bacteria are adept at aggregating enteric viruses on their surfaces^29,30,31^ and in the process stabilizing these virions and increasing their “resistance’ to inactivation under harsh conditions. Since the clarification process aggregates bacteria, it might consequently further aggregate enteric viruses which are already on bacterial surfaces. Taken together with the results of this study, it seems clear that WW clarification by size fractionation might be removing viruses from the filtrate and consequently the final concentrate may lose some virus diversity. This further emphasizes the need to ensure the virus is recovered from both FTS and filtrate for a more representative description of virus presence and diversity in any population.

### Seasonal variation

For all three viruses investigated in this study, we found a decrease in both virus presence and diversity between seasons 1 and 2 (Table 1, Figures 2, 4, 6, 7 and 8). This difference between seasons may be a result of the impact of nonpharmaceutical interventions used to mitigate the spread of SARS-CoV-2 in 2020/2021. Hence, virus type and variant diversity data generated from WW surveillance in this study was able to capture the impact of population-wide changes in human behavior in response to the SARS-CoV-2 pandemic.

It is however striking that this was also the case for CanPV. Whether this trend (a drop in variant detection and diversity) has been observed with domesticated animal infectious agents remains to be seen. It is however important to mention that while outside the USA, CanPV has been detected in dogs and red foxes^20,21,22,23^, in the USA it has only been detected in WW^24,25^. Hence, though CanPV is circulating in the US, we do not know its host(s) in the region. It has been shown that in household settings when humans have virus infections (like SARS-CoV-2 and monkeypox virus) they often transmit these to their dogs^32,33,34,35^. We do not know if the reverse happens with CanPV.

### CanPV genotyping

Though there had been previous reports ^20,21,22,23,24,25^ describing CanPV prior this study, there had been no attempt to genotype CanPVs. In this study, phylogenetic and pairwise identity analysis showed seven (7) distinct CanPV clusters with strong bootstrap support. Divergence within each cluster was less than 14% (Figure 5). We acknowledge that we do not know what these clusters represent with respect to the biology of CanPVs. It is, however, important to mention that the best-studied genus in subfamily *Ensavirinae* (supergroup 3) is *Enterovirus*. Within this genus, genotype designation was originally based on serotypes defined by neutralization assays. More recently, following studies^36,37^ demonstrating a correlation between serotypes and nucleotide sequence in the genomic region encoding capsid proteins, this has been replaced by pairwise identity in the capsid region and members of the same type have divergence ranging from less than 25% for Enterovirus A to around 12% for Rhinovirus B. Therefore, the less than 14% divergence observed in our CanPV lineages might be pointing to something fundamental in the biology of CanPV. Hence, here we refer to these lineages as *genotypes*, use them for classifying the CanPV variants and show that 1) there is ongoing circulation of multiple CanPV genotypes in the USA, 2) CanPV is circulating between continents, and 3) there are genotypes (like G3, G4 and G7) that are currently under-sampled (Table 3, Figure 5).

To facilitate our understanding of CanPV global diversity, dynamics, and geographical distribution, here we describe a new CanPV complete capsid amplification assay (Table S1, Figure 1). In this study, we show that the assay can be used for recovering the complete capsid sequence of CanPV from WW concentrates. We hypothesize that the workflow and assays described here should support the detection of CanPV in other sample types. Furthermore, we have improved the second-round assay previously described^24,25^ by adding a second one (Table S1, Figure 1). It is important to mention that both the ∼260bp and ∼975bp assays use the same forward primer but two different reverse primers. Hence, they can be coupled as a nested PCR protocol for CanPV detection in situations where long-range PCR reagents (as used here) are not available.

### EV diversity

The results of this study showed that EV-B was detected more often compared to other EV types. However, when considering variants, more EV-C variants were detected (Table 2). In fact, EV-C variants were more than 2x the EV-B variants detected (Table 2). This result suggests that the abundance of EV-C variants may not be due to assay bias, but rather reflective of virus dynamics in the population sampled. The EV-C abundance detected here is consistent with our previous findings^24,38^, but contradicts the prevailing paradigm that EV-Cs are not as common in North America^39,40,41^. We have previously shown that EV-C lineages were present in the USA that had not been sampled (or sequenced, but not made publicly available in GenBank) for around two decades^24^. This could be partly responsible for the paucity of EV-C types and variants circulating in the USA that are deposited in GenBank and consequently reflected in the dataset described in Brouwer and others^41^.

This narrative could also have its roots in the pervasiveness of cell culture-based virus isolation (and its biases) in EV history. Poliovirus (PV) is the best-studied EV (and EV-C) member and there are around 150 laboratories globally (as part of the global polio laboratory network [GPLN]) isolating PV on two cell lines; RD (human rhabdomyosarcoma) and L20B (transgenic mouse cell line expressing the human poliovirus receptor). It is important to note that while RD cell line is efficient at isolating PVs it does not do as well at isolating other non-polio EV-C members. It has been shown^4,42,43,44^ using both WW concentrates and fecal suspension that RD cell line has a bias for EV-B and when a sample containing both EV-B and nonPV EV-C members is inoculated into the cell line, it preferentially supports the replication of EV-B over EV-C. Furthermore, it has also been shown^45,46,47^ that samples considered negative for EVs because no isolate could be recovered on RD cell line, have an abundance of EV-Cs. Hence, studies showing an overabundance of EV-Bs^47,48,49,50^ in any population but in which EV isolation was based on RD cell line should be interpreted with caution as it might only be reflective of RD cell line bias for EVBs and not necessarily representative of EV diversity in such populations. How much cell culture bias has influenced the perception that EV-Bs are the most abundant EV type in the USA^39,40,41^ remains to be determined.

The emergence of circulating vaccine-derived polioviruses (cVDPVs), which are usually recombinants with nonstructural region of nonpolio EVC origin, is usually associated with low population immunity to polioviruses alongside vaccine poliovirus strains and circulating nonpolio EV-Cs^51^. The results of this study and our previous findings showing recombination between circulating EV-Cs in the USA^38^ indicate that if vaccine strains of poliovirus find their way into this population (as recently documented in June 2022 in a >20 years old man in Rockland County, New York, USA^52^), the use of inactivated poliovirus vaccine (IPV) suggests that cVDPVs can also emerge in this population, circulate and possibly seed other populations. Hence, monitoring EV-C dynamics in this population (and others globally where IPV is the mainstay of the vaccination effort) becomes critical as part of the eradication campaign.

### Ad41 diversity

In this study using WBE, we show that signatures of all three Ad41 hexon lineages recently associated with hepatitis-of-unknown-origin^27^ were present in the population sampled here during the study period (Table 4 and Figure 12). Our data also suggests that there might be circulating Ad41 variants (H2 and H4, Table 4) whose complete genomes have either not been sequenced or sequenced but not publicly available in GenBank. Hence, though as a DNA virus, Adenovirus evolution is slow^53^ when compared to RNA viruses, we demonstrate the feasibility of using WBE to monitor Ad41 diversity on a population scale. Specifically, our data shows that by targeting a protein under evolutionary pressure from the immune system (like the hexon), we can capture diversity (both amino acid substitutions [Table 4] and gross deletions like the 15aa deletion, Figure 11), and also use variant profiling coupled with CBS data to predict which lineages are likely present in the population at any point in time (Table 4 and Figure 8).

Analysis of Ad41 sequences publicly available in GenBank shows that the Xu and others^54^ assay used as our 2^nd^ round PCR assay in this study might be missing variants like those with the 135nt (i.e., 45aa) deletion in long-fiber shaft (Figure 11). The recurrence of large 45-135nt (15-45aa) deletions in the fiber shaft shows this genomic region might be too dynamic to use as target for Ad41 detection assays. Hence, it might be more beneficial to have detection assays target more stable Ad41 genomic regions.

While the tiled-amplicon approach described here shows that Ad41 surveillance using WBE is feasible, it also highlights some limitations. Analysis of Ad41 complete genome sequence data publicly available in GenBank showed that recombination is ongoing between the hexon gene and the fiber genes (Figure 10), which are greater than 8kb apart. Since the two amplification pools in the tiled amplicon approach are run in different tubes, there is no guarantee that the template genome(s) being amplified in both pools are from the same variant. Therefore, while data from the same contig (in this case ∼5kb) might be from the same template, it becomes difficult to ascertain whether overlapping tiles are from the same variant and consequently contiguous. It might therefore be necessary to implement long-range PCR assays that amplify penton-to-long-fiber or at least hexon-to-long-fiber and couple such to long-read sequencing strategies. Such an approach could enable scientists to study the co-evolution of distant Ad41 genomic regions using variants recovered from wastewater. It might be necessary to also consider this approach for WBE of other DNA viruses of clinical significance

### Limitations

In this study, we used samples that had been archived at -20°C. It is possible that the freeze-thawing process could have resulted in a reduction in virus titer. If association with bacteria protects particle integrity during the freeze-thaw process as it does to heat inactivation and bleach^31^, then this could be partly responsible for the larger recovery of EV type and variants in FTS. Furthermore, long-range PCR assays were used for our first round PCR assays. Hence, we might have missed fragmented genomes that might be detected by assays targeting smaller genomic regions or real-time RT-PCR assays. Finally, though we assayed both DNA and RNA viruses, we only screened for viruses with naked (non-enveloped) capsids because their particles are usually more stable in the environment and consequently more likely to have better-preserved genomes which is essential for our study as we amplified contigs ranging from 3.9-5kb. Studies are therefore needed replicating the experiments described here with fresh, unfrozen samples, which also target enveloped viruses.

## Methods

### Sample collection and processing

Samples archived at -20°C at the Human Health Observatory in Biodesign Institute, Arizona State University, Tempe, Arizona, USA, were used in this study. The study utilized 118 wastewater samples collected from ten different sites in two municipalities (population; ∼700,000) in Maricopa County, Arizona (USA) between October 2019 to March 2020 (Season 1) and October 2020 to March 2021 (Season 2) (12 month). Samples from all ten sites were from the same day of the month except for November 2020 when one site was collected within 24 hours of the others. Also, only eight (8) sites were collected in March 2020 because two of the locations were not sampled for logistic reasons due to the onset of the COVID-19 pandemic. Each sample was collected over 24 hours using time- or flow-weighted automated samplers.

For each of the 12 months, the ten samples per month were recovered from the freezer and thawed overnight. Subsequently, 200 ml of wastewater from each of the ten sites was filtered using ten 450nM membrane filters (Thermo Fisher Scientific, Waltham, MA, USA). The filtrates were pooled and concentrated to 2ml (∼1000x concentration) using 10,000 molecular weight (MW) cutoff centrifugal filters. The membrane filters were also recovered, and filter-trapped solids (FTSs) were resuspended by vortexing (Heidolph Instruments, Germany) for 10 minutes at 3000rpm in a 50ml centrifuge tube containing 25 ml of sterile PCR grade water containing 15 glass beads (3mm, Cole-Parmer, USA). After vortexing, the filters were removed, and the mixture was centrifuged for 20 minutes at 3900rpm and 4°C. The supernatant was recovered, pooled, and concentrated to 2 ml using 10,000 MW cutoff centrifugal filters. Hence, for each of the 12 months, there were two concentrates, one for filtrate and one for FTS.

### Nucleic acid extraction and polymerase chain reaction (PCR)

All 24 concentrates were subjected to nucleic acid extraction using the QIAamp viral RNA MiniKit following manufacturer’s instructions. The extract was used to screen for enteroviruses (EV), canine picornaviruses (CanPV) and adenovirus F41 (Ad41) using a collection of nested PCR assays (Table S1 and Figure 1). All assays were run using a BioRad 1000 thermal cycler (BioRad, Hercules, CA, USA). Amplicons were resolved on 2% agarose gels stained with GelRed (Biotium, Fremont, CA, USA) and viewed using BioRad Gel Doc XR+ system running Image lab 4.1 software with option to “highlight saturated pixels” enabled (BioRad, Hercules, CA, USA).

Since no CanPV complete capsid amplification assay existed at the time the study started, CanPV sequences publicly available in GenBank as of January 31, 2022, were downloaded and added to our local database, aligned and used to design primers for amplifying the complete capsid using Geneious Prime software^55^. The CanPV complete capsid amplification RT-PCR assay amplifies a ∼3900bp fragment of the genome encompassing the complete capsid protein coding genomic region. Amplicons from the CanPV complete capsid assay were used as template for two second round CanPV PCR assays targeting the VP2 (∼260bp) and VP2-VP3 (∼975 bp) gene segments. It is important to mention that both the ∼260 bp and ∼975 bp assays use the same forward primer, but two different reverse primers. Hence the region amplified by the ∼260 bp assay is the same as the first 260nt in the region amplified by the ∼975 bp assay. It is also crucial to emphasize that both assays are used independently as second-round assays in this study.

For Ad41, the complete genome was amplified in eight overlapping ∼5kb fragments in two multiplex assays (Table 1). Primers used in the Ad41 assay were designed using Primalscheme^56^. Ad41 complete genomes present in GenBank as of May 9, 2022, were downloaded, aligned and submitted to the Primalscheme web server with amplicon size set to 5,000bp. The eight overlapping amplicons were amplified in two multiplex assays. Pool 1 contained assays 1, 3, 5 and 7 while pool 2 contained assays 2, 4, 6 and 8. Please note that assays 7 and 8 both amplify the fiber genes.

### Sequencing

A random subset of amplicons generated from the second-round assays were cleaned and Sanger sequenced using their respective forward and reverse primers. This was to confirm that the first-round assays worked, and the target virus (and genomic region) was amplified. Subsequently, the first-round amplicons of all samples positive for the second-round assays were cleaned and used for library preparation and paired-end sequencing (2 × 250 bp) on an Illumina MiSeq sequencer (Illumina, San Diego, CA, USA) at the Biodesign Institute, Arizona State University, USA.

### Reads processing

The Illumina raw reads were processed on the KBase platform using default parameters^57^. Specifically, raw reads were trimmed using Trimmomatic v.0.36. The trimmed reads were then *de novo* assembled using metaSPAdes v3.15.3. Contigs were identified using Enterovirus genotyping tool^58^ and a BLASTn search of the GenBank database^59^.

For EVs and CanPVs, to confirm that variants found in both FTS and filtrate from the same WW sample were identical, trimmed reads from both fractions were merged and reassembled. Contigs that were present in the different fractions, but that coalesced in the merged analysis were considered the same.

For Ad41, trimmed raw reads were also mapped to the most similar complete genome from GenBank using the BBMap plugin in Geneious Prime^55^ with default parameters. Variant analysis was subsequently performed using the ‘find variant’ tool in Geneious Prime with default settings.

### Phylogenetics

Multiple sequence alignment (MSA) was done using ClustalW in MEGA X^60^, and Maximum-likelihood trees were constructed using 1,000 bootstrap replicates in IQ-Tree^61^. Prior to phylogenetic tree construction, the best-fitting nucleotide substitution model was selected using ModelFinder^62^. Pairwise identity was estimated using SDT v1.2.^63^ and heat mapper^64^.

### Structural modelling of Ad41

Structural modelling of Ad41 was done using ColabFold which implements Alphafold2 using MMseqs2^65^. Predicted structures were viewed and annotated using ChimeraX 1.4.^66^.

## Data Availability

The sequences described in this study have been deposited in SRA and GenBank under accession numbers PRJNA892620, OP643873 - OP643899 and OP627797 - OP627874.

## Data Availability

The sequences described in this study have been deposited in SRA and GenBank under accession numbers PRJNA892620, OP643873 – OP643899 and OP627797 – OP627874.

## Funding

The research reported in this publication was supported by the National Library of Medicine of the National Institutes of Health under Award Number U01LM013129 to RUH, MS and AV. The content is solely the responsibility of the authors and does not necessarily represent the official views of the National Institutes of Health.

## Acknowledgments

The authors thank the City of Tempe for sample collection. We also thank the Genomics Core at Biodesign Institute, Arizona State University for help with library preparation, Illumina and Sanger sequencing.

## Conflicts of Interest

R.U.H. is the founder of OneWaterOneHealth, a non-profit project of the Arizona State University Foundation. R.U.H. and E.M.D. are co-founders of AquaVitas, LLC, an ASU startup company operating in the field of wastewater-based epidemiology.

## Notes

### Summary of Updates

figure captions, minor grammar issue, and NCBI SRA and GenBank submission information

