## Supplemental Material for "Impact of sample clarification by size exclusion on virus detection and diversity in wastewater-based epidemiology"

**Table S1:** Reverse-transcriptase Polymerase chain reaction (RT-PCR) and PCR reaction conditions for assays used in this study. Note that all second round (nested) assays were done using GoTaq green PCR master mix. Ad41 assays 1 and 2 were done using Phusion green master mix. EV assays 1 and 2 and CanPV assay 1 were done using Superscript III Platinum Taq HiFi mix.

| Virus | | Region amplified | Amplicon size (bp) | RT | Pre-heat | # of cycles | Denaturation | Annealing | extension | Final incubation | References |
|---|---|---|---|---|---|---|---|---|---|---|---|
| EV | Assay 1 | Complete capsid 1 | ~3900 | 50ºC – 30 min | 94ºC – 2 min | 42x | 94ºC – 15 sec | 55ºC – 30 sec | 68ºC – 8 min | 68ºC – 5 min | Arita et al., 2016, Majumdar and Martin, 2018 |
| | Assay 2 | Complete capsid 2 | ~3900 | 50ºC – 30 min | 94ºC – 2 min | 42x | 94ºC – 15 sec | 55ºC – 30 sec | 68ºC – 8 min | 68ºC – 5 min | Majumdar and Martin, 2018 |
| | Assay 3 | Nested Partial VP1 | ~350 | N/A | 95ºC – 3 min | 35x | 95ºC – 30 sec | 55ºC – 30 sec | 60ºC – 30 sec | 72ºC – 5 min | Nix et al., 2006 |
| CanPV | Assay 1 | Complete capsid | ~3900 | 50ºC – 30 min | 94ºC – 2 min | 42x | 94ºC – 15 sec | 55ºC – 30 sec | 68ºC – 8 min | 68ºC – 5 min | This study |
| | Assay 2 | Nested partial VP2 | ~260 | N/A | 94ºC – 2 min | 35x | 94ºC – 15 sec | 55ºC – 30 sec | 60ºC – 30 sec | 68ºC – 5 min | This study |
| | Assay 3 | Nested partial VP2-VP3 | ~975 | N/A | 94ºC – 2 min | 35x | 94ºC – 15 sec | 55ºC – 30 sec | 60ºC – 60 sec | 68ºC – 5 min | Faleye et al., 2022 |
| Ad41 | Assay 1 | Complete Genome pool 1 | ~5000 for each of 4 non-overlapping amplicons | N/A | 94ºC – 3 min | 40x | 94ºC – 30 sec | 55ºC – 30 sec | 68ºC – 6 min | 68ºC – 10 min | This study |
| | Assay 2 | Complete Genome pool 2 | ~5000 for each of 4 non-overlapping amplicons | N/A | 94ºC – 3 min | 40x | 94ºC – 30 sec | 55ºC – 30 sec | 68ºC – 6 min | 68ºC – 10 min | This study |
| | Assay 3 | Nested Partial long fiber | ~600 | N/A | 95ºC – 3 min | 35x | 95ºC – 30 sec | 55ºC – 30 sec | 60ºC – 45 sec | 72ºC – 5 min | Xu et al., 2000 |

**Table S2:** Summary of Illumina raw reads generated, trimmed and mapped to EV contigs in this study. FTS = Filter-trapped-solids.

| | Month-Year | Conc-ID | Total # Raw reads | Total # trimmed reads | Total # mapped reads | Total # mapped reads (%) |
|---|---|---|---|---|---|---|
| Filtrate | Oct-19 | 1 | 2,050,970 | 2,026,030 | 458,197 | 22.62 |
| Filtrate | Nov-19 | 2 | 2,110,294 | 2,104,632 | 649,515 | 30.86 |
| Filtrate | Dec-19 | 3 | 2,594,172 | 2,582,252 | 594,354 | 23.02 |
| Filtrate | Jan-20 | 4 | 2,321,232 | 2,303,806 | 1,864,473 | 80.93 |
| Filtrate | Mar-20 | 6 | 2,035,078 | 2,019,214 | 1,325,652 | 65.65 |
| Filtrate | Dec-20 | 9 | 3,264,050 | 3,257,240 | 3,013,800 | 92.53 |
| Filtrate | Jan-21 | 10 | 1,920,390 | 1,914,528 | 1,055,898 | 55.15 |
| Filtrate | Feb-21 | 11 | 1,565,616 | 1,558,982 | 717,069 | 46 |
| FTS | Oct-19 | 13 | 1,632,010 | 1,627,374 | 959,548 | 58.96 |
| FTS | Nov-19 | 14 | 1,580,750 | 1,577,656 | 1,142,461 | 72.42 |
| FTS | Dec-19 | 15 | 2,492,324 | 2,484,688 | 1,617,641 | 65.1 |
| FTS | Jan-20 | 16 | 1,821,012 | 1,816,656 | 1,773,157 | 97.61 |
| FTS | Feb-20 | 17 | 1,363,620 | 1,359,258 | 1,298,427 | 95.52 |
| FTS | Mar-20 | 18 | 1,475,316 | 1,470,524 | 1,350,551 | 91.84 |
| FTS | Oct-20 | 19 | 1,863,568 | 1,858,834 | 621,050 | 33.41 |
| FTS | Nov-20 | 20 | 2,187,676 | 2,183,908 | 2,108,315 | 96.54 |
| FTS | Dec-20 | 21 | 1,485,720 | 1,482,590 | 1,418,636 | 95.69 |
| FTS | Jan-21 | 22 | 1,481,654 | 1,478,370 | 551,839 | 37.33 |
| FTS | Mar-21 | 24 | 1,463,408 | 1,458,982 | 1,147,286 | 78.64 |
| | | | 36,708,860 | 36,565,524 | 23,667,869 | 64.72 |

**Table S3:** Summary of Illumina raw reads generated, trimmed and mapped to CanPV contigs in this study. FTS = Filter-trapped-solids.

| | **Month-Year** | **Conc-ID** | **Total # Raw reads** | **Total # trimmed reads** | **Total # mapped reads** | **Total # mapped reads (%)** |
|---|---|---|---|---|---|---|
| Filtrate | Dec-19 | 3 | 930,978 | 929,244 | 630,397 | 67.84 |
| Filtrate | Jan-20 | 4 | 1,048,618 | 1,045,692 | 628,945 | 60.15 |
| Filtrate | Mar-20 | 6 | 1,021,308 | 1,018,062 | 618,284 | 60.73 |
| Filtrate | Dec-20 | 9 | 410,080 | 407,816 | 160,034 | 39.24 |
| Filtrate | Jan-21 | 10 | 890,000 | 887,056 | 523,488 | 59.01 |
| Filtrate | Mar-21 | 12 | 971,240 | 967,016 | 98,642 | 10.2 |
| FTS | Oct-19 | 13 | 1,146,120 | 1,143,542 | 801,737 | 70.11 |
| FTS | Nov-19 | 14 | 1,168,766 | 1,163,982 | 329,132 | 28.28 |
| FTS | Dec-19 | 15 | 1,000,374 | 998,860 | 913,946 | 91.5 |
| FTS | Jan-20 | 16 | 1,118,184 | 1,116,248 | 940,890 | 84.29 |
| FTS | Mar-20 | 18 | 1,671,848 | 1,668,836 | 1,498,275 | 89.78 |
| FTS | Nov-20 | 20 | 787,894 | 784,834 | 63,052 | 8.03 |
| FTS | Dec-20 | 21 | 826,726 | 823,142 | 658,421 | 79.99 |
| FTS | Jan-21 | 22 | 872,380 | 869,716 | 416,873 | 47.93 |
| | | Total | 13,864,516 | 13,824,046 | 8,282,116 | 59.91 |

**Table S4:** Summary of Illumina raw reads generated, trimmed and mapped to Ad41 in this study.

| | Month-Year | Conc-ID | Total # Raw reads | Total # trimmed reads | Total # mapped reads | Total # mapped reads (%) |
|---|---|---|---|---|---|---|
| Filtrate | Oct-19 | 1 | 1,949,546 | 1,939,546 | 1,016,632 | 52.42 |
| Filtrate | Nov-19 | 2 | 2,275,718 | 2,264,586 | 1,258,868 | 55.59 |
| Filtrate | Dec-19 | 3 | 2,090,046 | 2,080,078 | 1,461,773 | 70.27 |
| Filtrate | Jan-20 | 4 | 2,229,900 | 2,218,484 | 1,128,286 | 50.86 |
| Filtrate | Feb-20 | 5 | 1,997,038 | 1,982,070 | 993,865 | 50.14 |
| Filtrate | Mar-20 | 6 | 2,176,360 | 2,162,018 | 735,266 | 34.01 |
| Filtrate | Feb-21 | 11 | 2,193,634 | 2,179,436 | 165,956 | 7.61 |
| Filtrate | Mar-21 | 12 | 2,072,890 | 2,058,266 | 69,852 | 3.39 |
| FTS | Oct-19 | 13 | 2,235,250 | 2,222,254 | 1,307,496 | 58.84 |
| FTS | Nov-19 | 14 | 2,403,492 | 2,392,238 | 1,261,209 | 52.72 |
| FTS | Dec-19 | 15 | 2,430,600 | 2,417,524 | 1,608,144 | 66.52 |
| FTS | Jan-20 | 16 | 2,494,270 | 2,478,854 | 1,128,793 | 45.54 |
| FTS | Feb-20 | 17 | 2,039,716 | 2,029,004 | 1,297,690 | 63.96 |
| FTS | Mar-20 | 18 | 1,869,524 | 1,853,254 | 363,461 | 19.61 |
| FTS | | | 30,457,984 | 30,277,612 | 13,797,291 | 45.57 |

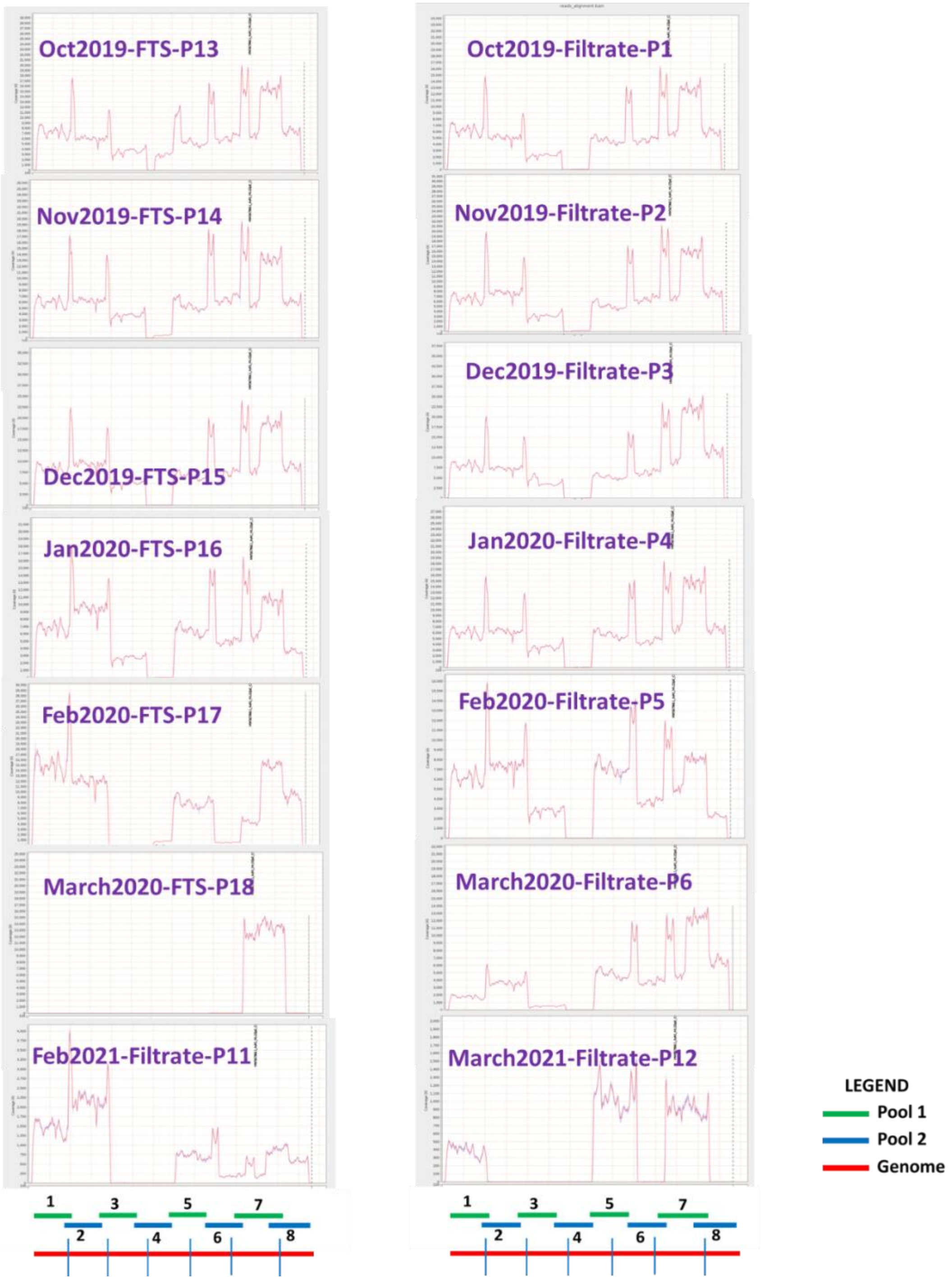


**Figure S1:** Mapped reads showing Ad41 genomic regions recovered by the complete genome assay. The schematic representation of the assay is shown below the mapped reads. Red line represents the genome divided into ~5kb blocks by the blue vertical lines. The green and blue lines are amplicons recovered from assays 1 and 2, respectively.
